# Chronological Mapping of Comorbidities in Alzheimer’s Disease and Vascular Dementia

**DOI:** 10.1101/2025.04.28.25326575

**Authors:** Chloe Walsh, Antigone Fogel, Ann-Kathrin Schalkamp, Vanessa Mabiala, Behnam Shariati, Cynthia Sandor, Mina Ryten, Ramin Nilforooshan, Payam Barnaghi

## Abstract

**Background:** Pre-existing long-term health conditions are highly prevalent in people living with dementia. There is an established relationship between these conditions, trajectories of decline and outcomes in dementia post-diagnosis. An understanding of when these conditions first occur and how they manifest over time within dementia populations remains unclear. The pathophysiology across major bodily systems changes before and after the clinical onset of dementia. However, the timing of various complex, chronic conditions, how they interact and their association with dementia are not yet fully understood. Understanding the complex relationship between multiple long-term conditions and dementia can enhance our understanding of the mechanisms of disease. This understanding can help identify “windows of opportunity” for early interventions in at-risk populations. Long-term conditions also continue to affect individual decline as the disease progresses. We set out to map these key comorbidities chronologically, specific to diagnoses of Alzheimer’s Disease and Vascular Dementia.

**Method:** We map comorbidities based on International Classification of Diseases (10th revision) system, in a population of people with Alzheimer’s Disease and Vascular Dementia from 20 years before through to 10 years after diagnosis. We used inpatient hospital electronic healthcare record data from 10,696 UK Biobank participants, with records spanning a median of 16.90 years (IQR = 10.10 years) for participants with dementia and 13.36 years (IQR = 13.52) for controls (no diagnosis of dementia). Controls were validated with lower polygenic risk scores than either dementia group.

**Results:** Characteristic comorbidity signatures were observed in individuals with Alzheimer’s Disease and Vascular Dementia. Up to 20 years before diagnosis, depressive episodes, osteoporosis, arthritis and irritable bowel syndrome are characteristic of Alzheimer’s Disease, but not Vascular Dementia or control cohorts. From 15 years before diagnosis, type 1 diabetes, functional intestinal disorder and chronic obstructive pulmonary disease begin to emerge. Up to 10 years before, symptoms involving fluid and food intake are uniquely associated with the cohort of people diagnosed with Alzheimer’s Disease. In Vascular Dementia, 20 years before diagnosis cerebral infarctions, type 1 diabetes, cerebrovascular disease, peripheral vascular disease, intestinal disorders and rheumatoid arthritis are early conditions in the Vascular Dementia cohort, not seen in control or Alzheimer’s Disease cohorts. 15 years before Vascular Dementia, an increase in mental health conditions emerge such as depressive episodes as well as rheumatoid arthritis. 10 years before Vascular Dementia, an additional burden of cerebrovascular diseases, hypotension and dorsalgia. From 5 years up to diagnosis of Vascular Dementia, intra-cerebral haemorrhage, mental and behavioural disorders due to tobacco begin to emerge within this population. In the years following Alzheimer’s Disease diagnosis, acute lower respiratory infections and skin conditions such as decubitus ulcer emerge up to 2 years after diagnosis. After a diagnosis of Vascular Dementia, pneumonitis and decubitus ulcer and pressure area, are unique to this cohort.

**Conclusion:** In this study, comorbid conditions were mapped in relation to Alzheimer’s Disease and Vascular Dementia pre- and post-diagnosis. Understanding prodromal and progressive changes in health over the course of these dementia sub-types could uncover opportunities for targeted screening, preventative measures and early interventions.

**Summary:** Dementia develops alongside multiple long-term health conditions^1–3^, yet the timing and subtype-specific patterns of these comorbidities remain unclear. Using hospital records from 10,696 UK Biobank participants, we mapped comorbidities from 20 years before to 10 years after diagnosis of Alzheimer’s Disease or Vascular Dementia.

We show that distinct comorbidity signatures precede each dementia subtype: depression, osteoporosis and intestinal disorders are early features of Alzheimer’s Disease, while Vascular Dementia is characterised by cerebrovascular and peripheral vascular diseases, type 1 diabetes and arthritis up to two decades before diagnosis. Post-diagnosis, conditions such as respiratory infections and pressure ulcers show divergent trajectories.

These findings reveal temporally distinct, disease-specific comorbidity profiles, suggesting differing pathological pathways and windows for targeted intervention. Understanding the longitudinal burden of comorbidities in dementia can inform screening, risk stratification, and preventative strategies long before clinical onset.

## Introduction

Comorbidities and long-term health conditions have a profound impact on the life of people with dementia^1,4^. Several decades before dementia is diagnosed, these long-term health conditions begin to manifest, increasing a person’s risk of developing dementia later in life. Following a dementia diagnosis, the impact of comorbidities becomes more complex and interrelated, contributing to decrease in quality of life and exerting a significant demand on health services^2,5^. Approximately half of the factors that influence susceptibility to dementia are unexplained by existing lifestyle, behavioural, socioeconomic or genetic factors^6^. The Lancet commission for dementia was a major study in the field, specifically addressing the risk factors of dementia for the purposes of prevention, intervention and care^7,8^. Long-term health conditions thought to increase the risk of dementia include untreated sensory impairments, metabolic disorders, hypertension, and depression^3^. There is emerging evidence that dementia could be considered as a systemic disease^9–11^. However, existing research either considers single or a sub-set of conditions and do not consider the temporal aspect of comorbidity patterns^12^.

Dementia comprises an umbrella of different sub-types, according to their most likely pathological cause. Examples include but are not limited to Alzheimer’s Disease, Vascular Dementia, Frontotemporal Dementia, Lewy Body Dementia and Parkinson’s Disease with Dementia^13^. There is evidence that dementia sub-types can be characterised by different comorbidities. A higher comorbidity load is associated with accelerated decline and mortality in people living with dementia^14^. Decline in cognitive ability and progression of dementia may be exacerbated by specific profiles of long-term health conditions and comorbidity burden^15,16^. Strong associations have also been established at a population level between comorbidity burden, types, and outcomes in people living with dementia^17–21^. Longitudinal analysis of dementia populations combined with multi-modal aspects of health have been investigated to identify early associated comorbidities but lacked a temporal aspect^22^. Those that included a temporal association showed that conditions related to hearing loss and the musculoskeletal system were associated with Alzheimer’s Disease (AD) up to five years before diagnosis, and in the same time span, cardiac and cerebrovascular diseases were associated with Vascular Dementia (VD); however, this work did not extend beyond dementia diagnosis and was conducted in a small population of 347 AD, 76 VD and 811 controls^23^. Many studies do not extend beyond diagnosis and only analyse the effects of comorbidities in isolation of other conditions or confounding factors. Interaction effects and a holistic view are necessary; the higher the comorbidity burden, the worse the outcome, particularly for those with dementia compared to controls. Using a more holistic approach, one study, although small, with a sample of 298 participants analysed the effects of self-reported (or relative/family reported) comorbidities, with International Classification of Disease - 10 (ICD-10) as a reference only, incorporating genetic risks such as apolipoprotein haplotypes, observing their impact on dementia outcomes^24^. Existing work investigated risk factors for dementia, using comorbidities as predictors of dementia incidence and outcomes including mortality^25,26^. Others incorporate comorbidities, biomarkers and other indications that can be used to identify individuals decades before developing dementia^27,28^. One study began forecasting individual progression and trajectories of disease, using a variety of methods from various longitudinal cohorts, however, did not span more than approximately 5 years^29^. A large-scale, data-driven approach, identifying long-term health conditions most associated with dementia sub-types, will lead to informed hypotheses for future dementia research and allow for more timely management for comorbidities preceding and following dementia diagnosis.

It remains unclear how the timing or temporal patterns of these conditions emerge and when they have greater influence or impact in relation to dementia. Earlier detection is important to identify key therapeutic avenues to meaningfully intervene and control the impact of these conditions on individuals and their association with dementia. Insights derived from temporal associations of conditions with dementia could further inform dementia research and support policymakers in developing pathways to identify individuals at risk of dementia earlier. Understanding common conditions that drive decline or emerge following a dementia diagnosis would also allow clinicians to better prepare for and manage these conditions. This ultimately can support individual patients’ quality of life and care. It is important that these insights be built upon existing resources to utilise the information already gathered within electronic healthcare records and systems. In this study, using electronic healthcare record data, we mapped the long-term health conditions of individuals with dementia diagnosed and documented by clinicians, spanning 20 years pre- and 10 years post-diagnosis, from the UK Biobank population database. We analysed a multitude of comorbidities over time to create characteristic comorbidity profiles in individuals with dementia sub-types AD and VD. This work observed how long-term health conditions changed and interacted over the course of 20+ years, for dementia and control (matched for age and sex) populations. We demonstrate the importance of these conditions in their association with dementia and post dementia diagnosis decline.

## Results

The UK Biobank population included 446,848 participants, with baseline assessments and in-patient ICD-10 codes available. Of 9,834 participants with potential dementia diagnoses, 50% (n=4,874) had a diagnosis of one dementia sub-type. 30% (n=2907) had two, 14% (n=1331) had three and 6% (n=722) had four or more; participants with four or more different dementia sub-type diagnoses were excluded (n=722). For each participant, an ‘Overall Mapping’ was established, as described in the Methods section (Study Design and Data Processing - Dementia Diagnosis). Participants diagnosed with Overall Mapping of either Unspecified Dementia (n=2114), Mild Cognitive Impairment (n=978), or Other dementia diagnosis (n=988) were excluded. The final cohort consisted of 5,348 people diagnosed with dementia, who were then matched to a control group (n=5,348) using age at diagnosis and sex.

In the final cohort of 5,348 participants with a dementia diagnosis, 3,775 had AD sub-type and 1,573 had VD sub-type. A control cohort of 5,348, who were matched to each dementia case based on age and sex, were included and defined as a population without a diagnosis of dementia in all of their in-patient data. This totalled 10,696 participants altogether. Of the 5,348 participants with dementia, 48% (n=2,545) were deceased, whereas in the control cohort, only 9% (n=498) were deceased. Data regarding deceased participants was last updated April 2024.

### Comorbidities associated with dementia diagnosis

A Mann-Whitney U Test was used to compare the long-term health conditions in those with dementia compared to their matched controls at the group level. For individual ICD-10 conditions, 264 unique conditions were significantly (*p*-value <0.05) associated with a dementia diagnosis. The top 20 of the most significant ICD-10 blocks are displayed in Figure 1, grouped by Overall Diagnosis. The full list of significant conditions is included in Supplementary Figure S1. Key comorbidities identified included circulatory and cerebrovascular disease, intestinal and urinary or renal conditions. In addition, other consequential complications were reported in the dementia cohort, including pneumonia, decubitus ulcers and pressure areas (bed sores), delirium and falls (*p*-value <0.05). The same analysis was conducted for block or group level diagnoses of the ICD-10 codes (Supplementary Figure S2). However, from this analysis alone, it was not possible to determine how these conditions influence one another or in what order they affect decline. To address this issue, we conducted a network analysis with a temporal aspect embedded.

**Figure 1:**
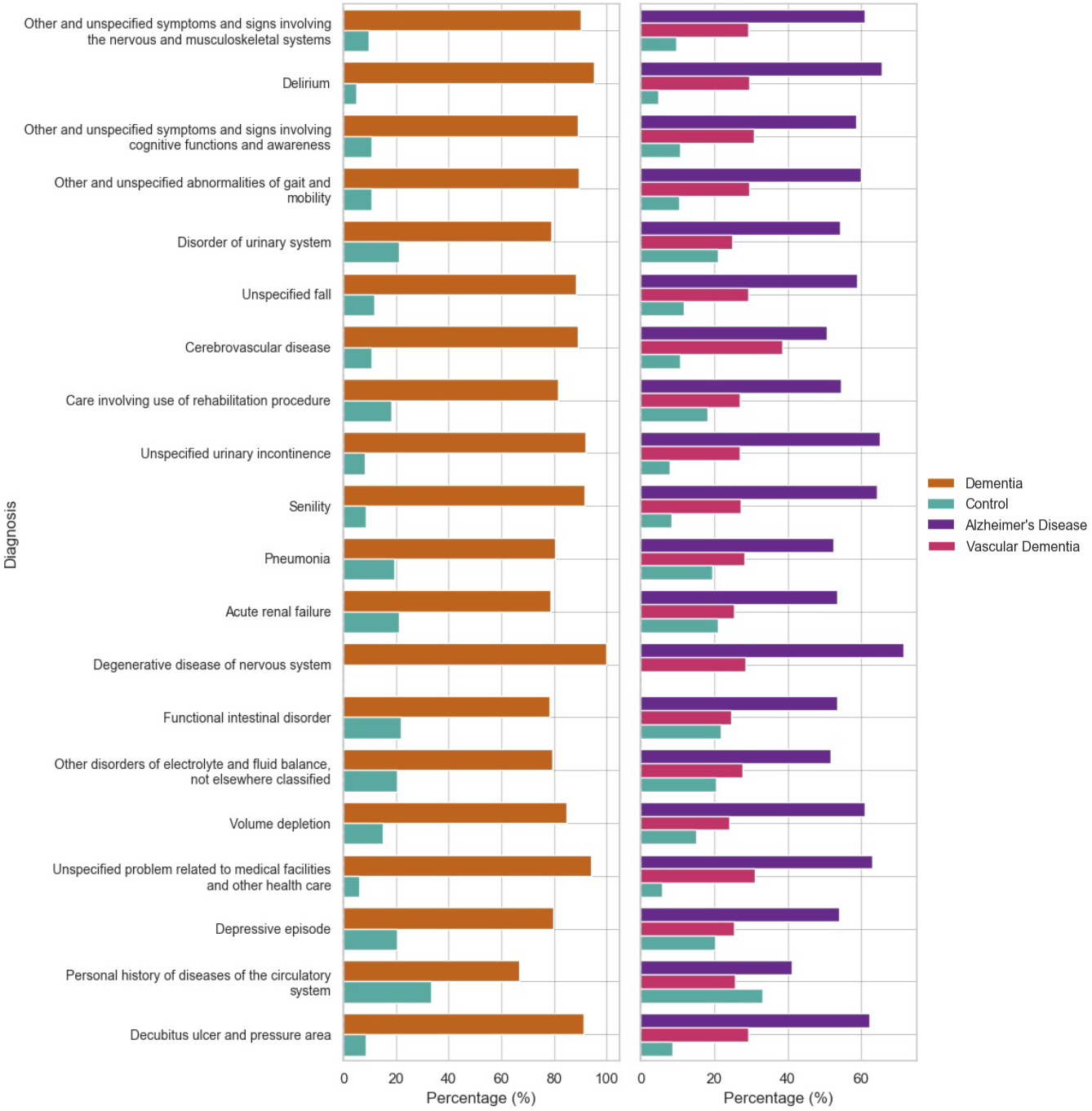
Prevalence of top 20 ICD-10 conditions that were significantly different between dementia and controls. Here, a Mann Whitney-U Test (all adjusted *p-value*s < 0.05, Bonferroni correction) was deployed, to compare association of each condition with each dementia subtype. The first panel (left) categorised by either controls or dementia cohort. The second panel (right) categorised by either controls or dementia subtypes Alzheimer’s Disease and Vascular Dementia. Conditions are ordered from top to bottom, in order of most to least significant. For readability purposes, conditions coded as “unspecified” had this removed however, they still correspond to the ICD0=-10 code. Comorbidities and complications are significantly associated with dementia including circulatory diseases, mental health episodes and complications such as infections.

### Control disease validation

To verify the findings that we had observed with AD and VD, we repeated the analysis using a control disease. This was identified as hip fractures, denoted by code ‘*S*72’ in ICD-10 coding. Supplementary Figure S3 shows the top 20 conditions that were identified as significantly associated with the cohort diagnosed with hip fractures, when compared with a population without hip fractures. Confirming our findings, these lists did not have any conditions in common with characteristic conditions of the AD or VD population. In addition, we analysed the prevalence of the conditions identified in Figure 1, in the cohort of people with hip fractures. Supplementary Figure S4 details the proportion of people with hip fractures, that were diagnosed with the top 20 most significant conditions associated with a dementia diagnosis of either AD or VD, as seen in the dementia cohorts (Figure 1).

### Characteristic comorbidity analysis of Alzheimer’s Disease and Vascular Dementia

Considering dementia as a disease with multiple systemic contributions and effects, there is an unmet need to provide a comprehensive overview of the timing of comorbid conditions across the life span and analysis of how they interact. To address this challenge, we utilised the comorbidities that were most associated or characteristic to each dementia sub-type, AD and VD, obtained from the analysis described above (Figure 1). A summary of these conditions is shown in Figure 3.

To first establish the association of individual comorbidities with either AD or VD, a logistic regression model was used. Metrics relating to the model performance are detailed in Supplementary Tables S1 for AD and S2 for VD; only conditions diagnosed before the dementia diagnosis were used in each model. The corresponding odds ratios and confidence intervals of each condition, with respect to the outcome of either AD or VD, adjusted for sex and age at diagnosis, are shown in Figure 2a and b. This figure only presents the conditions that were found to have a significant (*p*-value <0.05) association with either AD or VD. Each condition and corresponding odds ratio for each dementia sub-type are displayed in Supplementary Table S3 and S4. In AD, conditions that were most associated with a diagnosis of AD included symptoms and signs concerning fluid and food intake (*p-value* <0.001), 10-7 years before and delirium (*p-value* = 0.001), 5-2 years before. However, for VD, cerebrovascular disease (*p-value* = < 0.001), 15-10 years before and intracerebral haemorrhage (*p-value* < 0.001), 7-5 years before were most associated with this cohort. A detailed breakdown of the regression analysis is described below, within this section.

**Figure 2:**
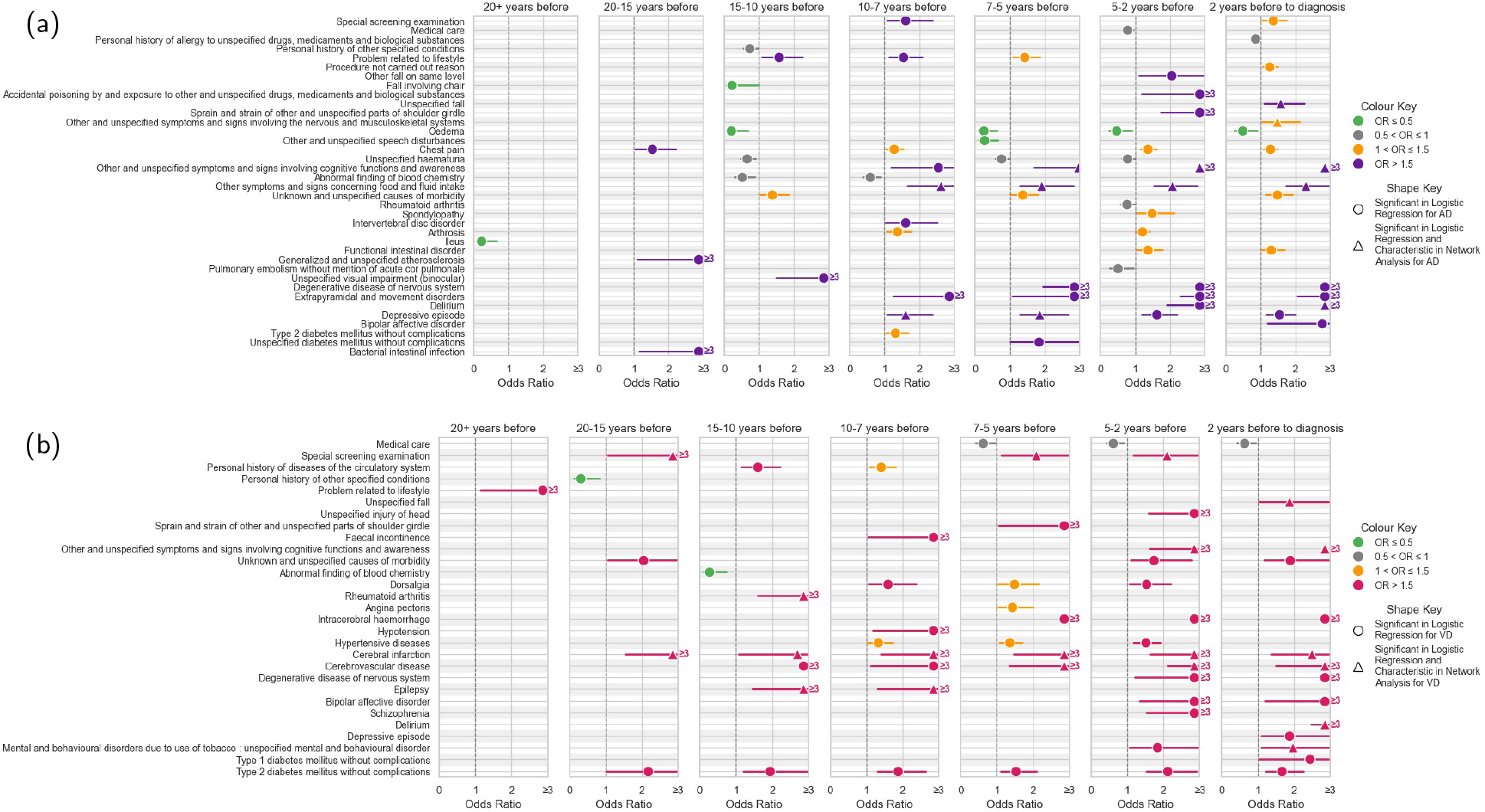
Significantly associated conditions and their odds ratios (with confidence intervals) for diagnosis of (a) Alzheimer’s Disease and (b) Vascular Dementia, adjusted for sex and age at dementia diagnosis. The logistic regression model included only conditions diagnosed prior to dementia diagnosis that featured as significantly associated with dementia cohort, in Mann Whitney-U tests. This figure displays only conditions from the logistic regression model, with a significant association (*p-value* < 0.05) with each dementia subtype. Data points refer to odds ratio value and range of 95% confidence intervals. Depressive episodes, and complications related to food or fluid intake show increasing association with Alzheimer’s Disease from 20 years before diagnosis up to the point of diagnosis. Cerebral infarctions and cerebrovascular disease show consistently strong associations with Vascular Dementia from 20 years before diagnosis up to the point of diagnosis. Odds ratio values of greater than three are plotted as ‘* 3’. (AD: Alzheimer’s Disease).

**Figure 3:**
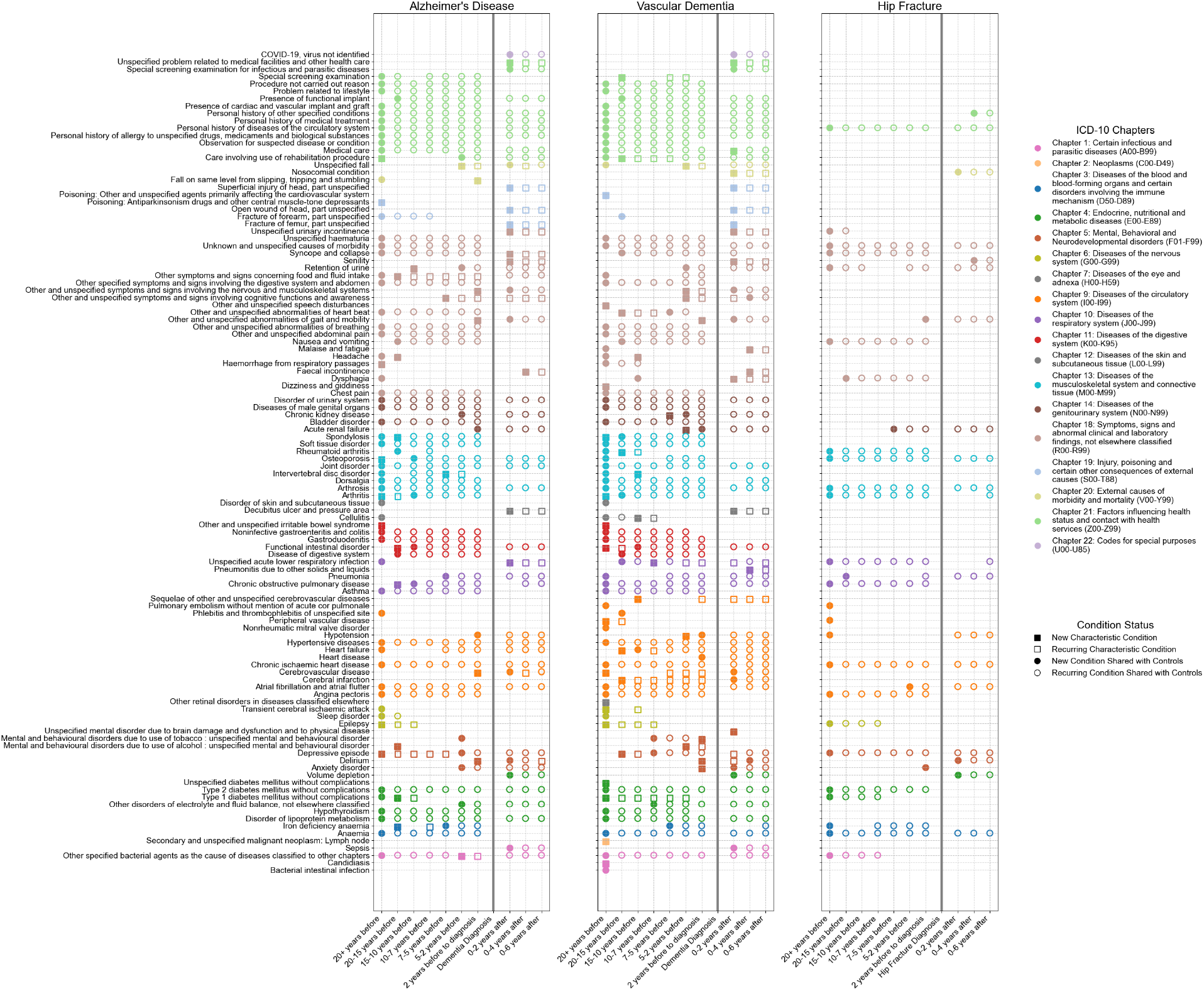
Chronological mapping of comorbidities associated with Alzheimer’s Disease (left panel), Vascular Demen- tia (centre panel) and Hip Fractures as the Control Disease (right panel), from 20 years before diagnosis to 6 years after. Each time frame is shown, with the timing of dementia diagnosis (or hip fracture diagnosis) marked by a solid vertical grey line. The conditions displayed represent individual types of ICD-10 coded diagnoses. As detailed in the figure legend (‘Condition Status’), squares represent comorbidities characteristic of the cohort at the given time frame, not seen in controls, while circles indicate comorbidities shared between the cohort and matched controls. Filled shapes denote new comorbidities, whereas unfilled shapes represent reoccurring conditions. Colours correspond to higher-level ICD-10 Chapters, coded in the ‘ICD-10 Chapters’ legend. None of the conditions found to be characteristic of Alzheimer’s Disease or Vascular Dementia, were characteristic of the cohort with hip fractures although, they shared some conditions with their matched controls, often regarded characteristic of dementia cohorts. For example, depressive episodes characteristic for both dementia subtype cohorts, were not characteristic of the cohort with hip fractures. (ICD: International Classification of Disease)

Although there were fluctuations in odds ratio values across the different cohorts, some trends can be observed for each subtype. For AD subtype, odds ratio measures for conditions such as depressive episodes, delirium and extrapyramidal and movement disorders increased as the time frames got closer to diagnosis (Figure 2a). However, conditions such as arthrosis showed a reduction in the odds ratio across the time frames. For VD, hypertensive diseases, intracerebral haemorrhage and type 2 diabetes all showed an increase in odds ratio across the time frames, as we get closer to the diagnosis. Reduction in odds ratios for epilepsy were observed in the VD cohort (Figure 2b).

Here we provide a more detailed description of the the regression analysis for AD and VD cohorts. In the cohort for AD, 10-7 years before diagnosis, symptoms and signs concerning fluid and food intake (Odds Ratio: 2.61, 95% CI: 1.64-4.15, *p-value* <0.001), extrapyramidal and movement disorders (Odds Ratio: 3.05, 95% CI:1.13-8.25, *p-value* = 0.028), arthrosis (Odds Ratio: 1.37, 95% CI: 1.06-1.76, *p-value* = 0.015), intervertebral disc disorder (Odds Ratio: 1.59, 95% CI: 1.01-2.50, *p-value* = 0.045), depressive episodes (Odds Ratio: 1.60, 95% CI:1.05-2.44, *p-value* = 0.03) and type 2 diabetes (Odds Ratio: 1.31, 95% CI:1.02-1.68, *p-value* = 0.032) were characteristic (Figure 2a). Both fluid and food intake symptoms as well as depressive episodes continued to be strongly associated with AD, until diagnosis. From 5-2 years before AD, symptoms involving cognitive functions and awareness were strongly associated with AD (Odds Ratio: 4.72, 95% CI:3.12-7.13, *p-value* < 0.001), as well as delirium (Odds Ratio: 4.10, 95% CI:1.78-9.48, *p-value* = 0.001). Within 2 years of AD diagnosis, bipolar affective disorder became strongly associated with this cohort (Odds Ratio: 2.76, 95% CI:1.16-6.55, *p-value* = 0.021).

For the VD cohort, from 20-15 years before diagnosis, metabolic conditions including type 2 diabetes were strongly associated with a VD diagnosis (Odds Ratio:2.16, 95% CI:1.01-4.63, *p-value* = 0.047) (Figure 2b). There is a sustained and consistent association of type 2 diabetes with VD until diagnosis. At 20-15 years before, cerebral infarctions (Odds Ratio:5.01, 95% CI:1.64-15.30, *p-value* = 0.005) and disorders of the urinary system (Odds Ratio:2.35, 95% CI:1.00-5.49, *p-value* = 0.049) all have strong associations with VD (Figure 2b). Cerebral infarctions remain consistently significantly associated with VD, until diagnosis. From 15-10 years before VD diagnosis, cerebrovascular disease (Odds Ratio:13.90, 95% CI:3.45-56.00, *p-value* = < 0.001), epilepsy (Odds Ratio:4.20, 95% CI:1.53-11.49, *p-value* = 0.005) and rheumatoid arthritis (Odds Ratio:4.31, 95% CI:1.61-11.60, *p-value* = 0.004) were strongly associated (Figure 2b). Cerebrovascular disease remained significantly associated with VD until diagnosis, consistently at each time frame.

Up to 10 years before VD, dorsalgia (Odds Ratio:1.58, 95% CI:1.04-2.40, *p-value* = 0.034), hypertensive diseases (Odds Ratio:1.33, 95% CI:1.03-1.72, *p-value* = 0.029) and hypotension (Odds Ratio:3.01, 95% CI:1.18-7.68, *p-value* = 0.021) were significantly associated with VD (Figure 2b). Other symptoms of note included personal history of diseases of the circulatory system (Odds Ratio:1.40, 95% CI:1.08-1.81, *p-value* = 0.011) (Figure 2b). From 7-5 years before VD, intracerebral haemorrhage (Odds Ratio:8.41, 95% CI:3.41-20.78, *p-value* < 0.001) emerges as strongly associated with a VD diagnosis (Figure 2b). Within 5 years of VD diagnosis mental and behavioural disorders due to use of tobacco (Odds Ratio:1.83, 95% CI:1.06-3.16, *p-value* = 0.03) bipolar affective disorder (Odds Ratio:3.20, 95% CI:1.32-7.73, *p-value* = 0.01) and schizophrenia (Odds Ratio:5.90, 95% CI:1.64-6.90, *p-value* = 0.001) and injuries of the head (Odds Ratio:7.42, 95% CI:1.67-32.84, *p-value* = 0.008) were all strongly associated with this cohort (Figure 2b). Symptoms and signs involving cognitive functions and awareness emerge (Odds Ratio:3.37, 95% CI:1.64-6.89, *p-value* = 0.001) up to 5 years before VD diagnosis. Mental and behavioural disorders due to the use of tobacco and bipolar affective disorder remain significantly associated with VD until diagnosis (Figure 2b). Up to 2 years before VD, depressive episodes (Odds Ratio:1.87, 95% CI:1.14-3.07, *p-value* = 0.013), type 1 diabetes (Odds Ratio:2.44, 95% CI:1.02-5.82, *p-value* = 0.045) and falls (Odds Ratio:1.87, 95% CI:1.04-3.36, *p-value* = 0.036) were strongly associated with this cohort, much later than what was observed in the AD cohort.

### Undirected network analysis of comorbidities associated with Alzheimer’s Disease

To consider the interactions between comorbidities and their wider importance in the context of overall health and rate of decline, for each dementia sub-type, undirected network analysis was applied per time frame for both dementia and control cohorts (Figure 3). As an example, we have displayed the corresponding network analysis results from 15-10 years before for AD and VD (Supplementary Figure S6a and b). In the context of this work, networks demonstrated the importance of each condition relative to all other conditions, which were quantified using the degree centrality of the networks. The same analysis was conducted for block or group level diagnoses of the ICD-10 codes (Supplementary Figure S5). For each time frame, the top twenty conditions according to centrality are displayed in Supplementary Figure S7 for AD and Supplementary Figure S8 for VD. Per time frame, a centrality measure closer to 1.0 indicates that the condition is central to that network; for AD, conditions such as heart, circulatory and hypertensive diseases were some of the first to increase in centrality metric, to within 0.5-1.0, approximately 15-10 years before diagnosis (Supplementary Figure S7). After diagnosis, complications, including delirium, emerge as highly important conditions for each network. With VD, a similar pattern to AD is observed with circulatory and heart diseases as important central conditions after diagnosis (Supplementary Figure S8). However, hypertensive diseases did not emerge as early as they did in the AD cohort. After VD diagnosis, no notable new conditions, not seen in AD, emerged as centrally important.

From the network analysis, we were able to establish conditions that were characteristic of AD or VD, per time frame (Figure 3). New conditions diagnosed 20 years before AD included mental health conditions such as depressive episodes. Neurological conditions such as epilepsy were characteristic of the AD cohort at this time (Figure 3). These were accompanied by other chronic conditions, including unspecified irritable bowel syndrome, arthritis and osteoporosis. Many different reports of in-patient results, signs and symptoms were reported at 20 years before AD diagnosis including: haemorrhage from respiratory passages and report of ‘poisoning’ from medications used for central muscle-tone depressants, often covering overdoses or misuse of the medication (Figure 3). Interactions with healthcare services at 20 years before diagnosis of AD included care and rehabilitation procedures, as one would expect from in-patient hospital patients receiving care packages. As we move closer to AD diagnosis, from 15 years before (Figure 3), other long-term health conditions including iron deficiency anaemia, type 1 diabetes, alcohol-induced mental and behavioural disorders, chronic obstructive pulmonary disease, functional intestinal disorders and spondylosis begin to be diagnosed and identified, characteristic of the AD cohort (Figure 3). Symptoms range from headache to symptoms concerning food and fluid intake. From 15 to 7 years up to diagnosis of AD, symptoms such as retention of urine were the only new, unique symptoms for the AD cohort. Up to 5 years before AD diagnosis, invertebral disc disorder was the only new, unique condition associated with the AD cohort and signs of loss of cognitive functions and awareness at this time (Figure 3). Up to 2 years before diagnosis, falls and bacterial agents as cause of diseases emerged, followed by falls from slipping/tripping/stumbling in the remaining 2 years before AD (Figure 3). Symptoms identified closest to AD include nervous system and musculoskeletal symptoms as well as symptoms related to abnormality of gait. At this time frame, cerebrovascular diseases and delirium were also characteristic of the AD cohort, compared to controls; however, a similar pattern was observed with VD. Symptoms involving concern of fluid and food intake as well as depressive episodes were seen to re-occur over at least five different time frames (Figure 3). Most other conditions were then shared amongst the age- and sex-matched control cohorts.

Once the AD diagnosis was made, 10 new diagnoses were identified as characteristic of the AD cohort after diagnosis (Figure 3). In the years following AD diagnosis, acute lower respiratory infections and bed sore incidence occur consistently up to 6 years after diagnosis. Other complications, including urinary and faecal incontinence, senility, syncope and collapse, and a range of injuries, including head wounds and fractures, were characteristic of the AD cohort (Figure 3).

### Undirected network analysis of comorbidities associated with Vascular Dementia

Compared with AD, the VD cohort showed a more unique profile (Figure 3). In VD, 20 years before diagnosis bacterial infections, malignant neoplasms, type 1 diabetes (and unspecified diabetes), epilepsy, transient cerebral ischaemic attack, retinal disorders occur; cerebrovascular disease, peripheral vascular disease, functional intestinal disorders, irritable bowel syndrome, arthritis and spondylosis were characteristic of the VD cohort (Figure 3). 15 years before diagnosis of VD, depressive episodes, cerebral infarction, heart failure and rheumatoid arthritis emerged. 10 years before VD, additional cerebrovascular diseases, cellulitis and intervertebral disc disorders occurred. Combining time frames, from 7 years up to diagnosis of VD, acute lower respiratory infections, chronic kidney disease, mental and behavioural disorders due to tobacco and alcohol and hypotension all begin to emerge. For VD, the conditions identified as unique were diagnosed earlier compared with AD (Figure 3). Conditions characteristic of this cohort were associated across at least six different time frames in advance of a VD diagnosis, including type 1 diabetes and cerebral infarction. Comorbidities that precede VD diagnosis were more specific to this cohort than in age- and sex-matched controls.

In the cohort with VD, there were 14 new diagnoses that were characteristic of this cohort after diagnosis was made (Figure 3). Brain damage and dysfunction due to physical disease, pneumonitis and bed sores were characteristic of the VD cohort. Other complications include dysphagia, faecal and urinary incontinence, malaise and fatigue, senility, wounds of the head and other fractures (Figure 3).

Combining results from a multi-level approach, of statistical and analytical techniques including logistic regression and Bayesian network analysis, we created a list for AD, for both before and after diagnosis including: ‘Other symptoms and signs concerning food and fluid intake’, ‘Other symptoms and signs involving cognitive functions and awareness’, ‘Depressive episode’, ‘Intervertebral disc disorder’, ‘Delirium’, ‘Functional intestinal disorder, unspecified’, ‘Unspecified fall’, ‘Other symptoms and signs involving the nervous and musculoskeletal systems’. For VD, this combined list includes: ‘Cerebral infarction’, ‘Care involving use of rehabilitation procedure’, ‘Cerebrovascular disease’, ‘Rheumatoid arthritis’, ‘Epilepsy’, ‘Hypotension’, ‘Other and unspecified symptoms and signs involving cognitive functions and awareness’, ‘Mental and behavioural disorders due to use of tobacco’, ‘Delirium’, ‘Depressive episode’, ‘Falls’, ‘Type 1 diabetes mellitus without complications’, ‘Other abnormalities of gait and mobility’.

### Control disease validation

To validate the patterns established using network analysis and the characteristic patterns associated with AD and VD, we repeated the analysis with the control disease population of people with hip fractures. When compared with their matched controls, the population of people with hip fractures shared many of the same characteristic comorbidities identified in AD and VD however, none of the conditions listed in the figure were identified as characteristic to hip fracture populations (Figure 3). This further validated the characteristic patterns that were identified in AD and VD cohorts.

### Outcomes post-diagnosis of Alzheimer’s Disease and Vascular Dementia

The impact of comorbidity profiles, including types and numbers of conditions, can affect outcomes in populations with dementia compared to controls. As a measure of an outcome, we used the time to death to establish how the type of dementia affects survival rates compared to controls. To observe an overall group effect, a survival analysis of dementia and the control cohort revealed a faster decline in those diagnosed with dementia compared to control counterparts, matched on sex and age at diagnosis (Figure 4).

**Figure 4:**
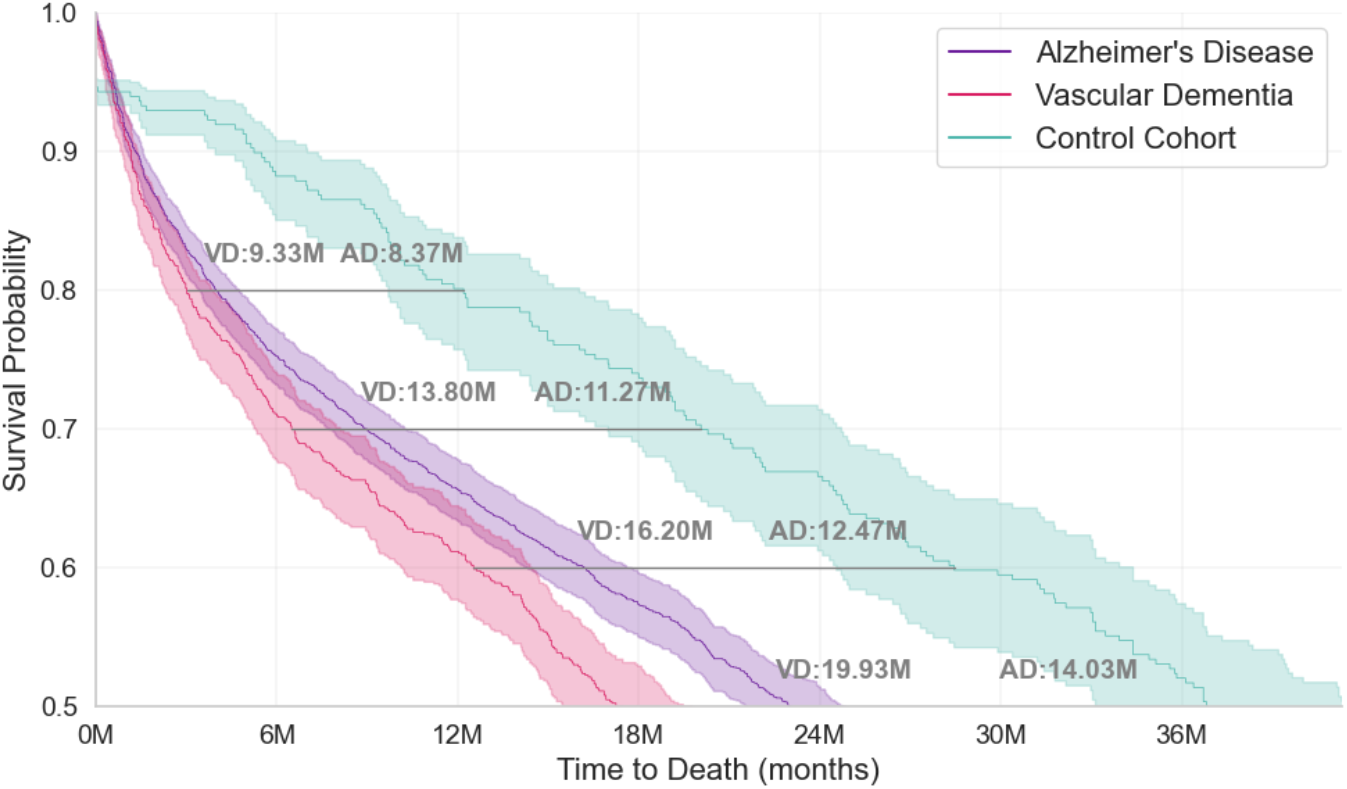
Kaplan-Meier Survival analysis for Alzheimer’s Disease and Vascular Dementia vs. controls. Annotations show how much earlier (in months) the Alzheimer’s Disease and Vascular Dementia cohorts reached corresponding survival probabilities compared to controls, up to 0.5 survival probability. All controls were matched based on sex and age at diagnosis of dementia. Vascular Dementia cohort showed the fastest decline overall, with more lag behind controls than the Alzheimer’s Disease cohort.(AD: Alzheimer’s Disease, VD: Vascular Dementia).

The analysis was limited to 67 months after dementia diagnosis. The lag was calculated to quantify the time difference in survival outcomes between the two groups, demonstrating how much earlier the dementia group reaches equivalent survival probabilities (Figure 4). Only survival probabilities up to 0.5 shown here, remaining results in Supplementary Table S6. From the point of diagnosis of dementia to the first six months post-diagnosis, those without dementia survive approximately 11 months longer than people with an AD diagnosis and approximately 14 months longer than those with VD. This pattern is consistent through to death, with the population with VD declining faster than those with AD.

Demographic data for dementia cohorts are detailed in Figure 5. A breakdown of the cohort’s ethnicity by dementia subtype is detailed in Supplementary Table S5. The ICD-10 diagnosis records from in-patient hospital data spanned a median of 16.90 years (IQR = 10.01 years) for the dementia cohort, while the control cohort records spanned a median of 13.36 years (IQR = 13.52 years). In addition, both dementia subtype groups had, on average, significantly (adjusted *p-value* = < 0.001) higher polygenic risk scores compared with the matched control cohort (Figure 5c). When comparing within subtypes, both AD and VD showed significantly higher (adjusted *p-value* = < 0.001) polygenic risk scores compared with matched controls. Across dementia subtypes, AD group showed significantly higher (adjusted *p-value* = < 0.001), compared with VD group. Overall, there were more comorbidities diagnosed in the dementia group when compared with controls (Figure 5d).

**Figure 5:**
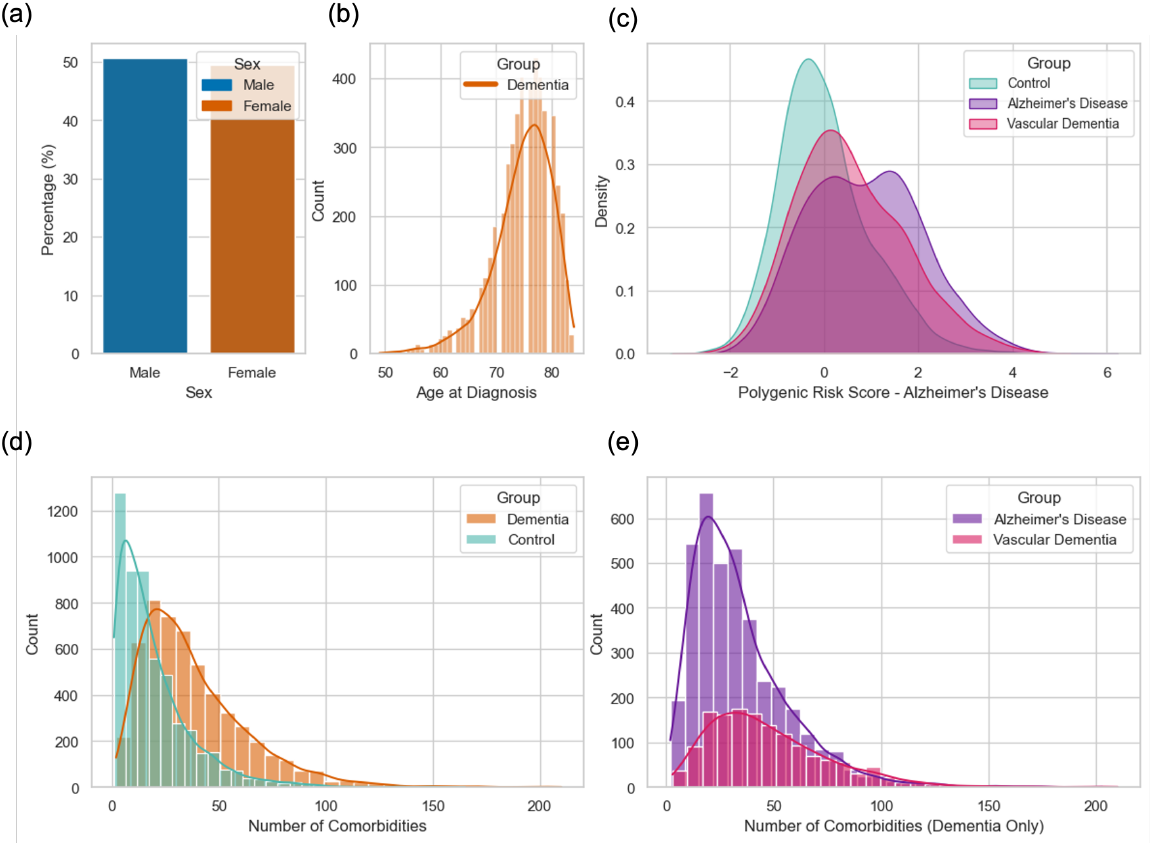
Baseline characteristics of dementia subtypes and control cohorts. (a) Percentage distribution of female and male participants, for dementia and control groups. (b) Distribution of age at diagnosis, for dementia cohort. (c) Polygenic risk scores for Alzheimer’s Disease based on each sub-type group. The Alzheimer’s Disease and Vascular Dementia subtype groups had significantly (adjusted *p-value* = < 0.001) higher polygenic risk scores than the controls. (d) Distribution of number of diagnosed comorbidities per group for dementia and control cohorts. Dementia cohort showed on average, higher numbers of diagnosed comorbidities as compared with control cohort. (e) Distribution of number of diagnosed comorbidities per group for dementia cohort only, grouped into each sub-type, Alzheimer’s Disease and Vascular Dementia.

In summary, we observed unique, temporal comorbidity profiles associated with AD and VD. These profiles were specific to time frames before and after diagnosis, compared with sex- and age-matched controls. Logistic regression established how much each condition was individually associated with either AD or VD. The odds ratio measures indicated how strongly each condition was associated. The centrality measures of the networks indicated the importance of conditions central to each network, enabling us to identify key therapeutic clinical targets. Mapping these conditions across time frames allows visual mapping of these unique comorbidity profiles. Up to 15 years before diagnosis, depressive episodes, epilepsy, functional intestinal disorder, spondylosis and iron deficiency anaemia were most strongly associated with an outcome of AD diagnosis. However, cerebral infarction, cerebrovascular and peripheral vascular disease, heart failure, type 1 diabetes, mental and behavioural disorders due to tobacco use, epilepsy, functional intestinal disorder, rheumatoid arthritis, spondylosis and chronic kidney disease were more distinctly associated with a VD diagnosis. The comorbidity landscape shifts after diagnosis of either AD or VD. Up to four years after diagnosis, bacterial/viral infections, renal failure and injuries to the head were characteristic of both dementia sub-types.

## Discussion

We established a chronological mapping of comorbidities across the lifespan in the context of AD and VD, compared with age- and sex-matched control populations. In a large cohort of 10,508 people in the UK, we systematically traced temporal patterns of long-term health conditions using data from in-patient electronic health records over a period of 20 years before to 10 years after AD or VD diagnosis. Many current observational studies lack a standardised method for categorising individuals into specific dementia sub-types, often lacking a definition of how they identified populations of people with dementia or what sub-types were included. We ensured that our approach to identify dementia populations and sub-types could easily be reproduced and replicated in other population datasets. Comorbidities that showed a higher statistical association with AD, prior to diagnosis, included depressive episodes and issues with fluid and food intake up to 10 years before diagnosis, while falls and delirium became prominent closer to diagnosis. In contrast, VD showed a distinct association, with circulatory diseases such as cerebral infarctions occurring up to 20 years prior and rheumatoid arthritis and epilepsy emerging 15 years before diagnosis. Up to 7 years before VD, cerebrovascular diseases were characteristic, and within two years, mental disorders and delirium became evident. Using statistical modelling techniques, our methods used existing populations to identify common pathways and long-term health conditions associated with either AD or VD. Models such as logistic regression consider the conditions and their association with the outcomes in isolation while confounding for age at diagnosis and sex. Interactions between features at previous or similar time frames cannot be inferred using these methods. To address this issue, we extended the analysis using a network analysis, with a temporal aspect embedded. Dementia care policies in the UK and around the world prioritise prevention and leveraging existing national healthcare resources^30^. This work builds on these principles, utilising existing electronic healthcare record data to track comorbidities preceding AD and VD diagnoses from longitudinal data collected as part of routine healthcare services. We identified critical “windows of opportunity” for potential interventions, tailored to each sub-type. This work complements existing approaches to dementia risk screening and informs targeted clinical practice strategies.

### Incidence of comorbidities may act as a flag for disease exacerbation

Some long-term health conditions identified in this study, including diabetes mellitus, are usually diagnosed earlier in a lifespan. Importantly, VD cohort showed an earlier and more consistent association with type 1 diabetes compared with AD. The VD cohort also showed additional metabolic disorders that were characteristic of VD but not AD. Broadly, both types of diabetes mellitus have been shown to be associated with increased cognitive dysfunction and dementia^31^. In this study, we identified a specific window at which type 1 diabetes was considered characteristic of the VD cohort, from 20 years through to diagnosis of VD, not seen in the AD cohort. Further work with comparison across different electronic healthcare records in both primary and secondary care settings would increase fidelity of these findings. Worsening glycaemic control has been associated with a greater risk of dementia^32^.

Exclusive to the VD cohort, symptoms of changes in gait and mobility were characteristic. Mobility decline has been a component of decline that is shown to vary depending on dementia sub-type; however, a longitudinal study of 766 older adults found that VD exhibited the fastest rate of decline^33^. At the point of conversion to VD, a longitudinal study over five years found that quantifying changes in gait are predictive of decline, with the pace of gait an important predictor of VD^34^. Changes and measures of mobility and gait are easily quantifiable but, importantly, cheaper than alternative assessments of decline such as imaging measures; nonetheless, measures such as mobility and gait serve as equally important early indicators of decline^35^.

We take a combined approach to temporal patterns of comorbidities, addressing how they occur and interact as a whole, not in isolation from one another. Interaction effects are important to consider, depending on the time frame in which they occur. Decades before onset of AD, it has been shown that interaction of intestinal disorders and clinical psychiatric symptoms are highly interrelated^36^. Other intestinal diseases including inflammatory bowel disease, along with inflammatory-related mechanisms have also been strongly associated with an increased risk of AD, in national longitudinal cohorts, both cross-sectional and case-control^37,38^. When diseases are modelled independent of their relation to dementia, in this UK Biobank cohort, there are clear links between cardiovascular comorbidities and type 2 diabetes, the same pattern we observed here^39^. Rheumatoid arthritis, as an autoimmune condition, has been associated with dementia, particularly VD^40^. To further support the hypothesis regarding autoimmune conditions and their associations with dementia, anti-inflammatory medications have been suggested as potential therapeutic targets for dementia, with varying degrees of results and outcomes^41–45^. This complex relationship between autoimmune conditions and their association with dementia is reflected in this cohort.

Characteristic to AD, from early on, were changes in food and fluid intake, consistently, not observed in the VD cohort. In AD, changes in eating and drinking behaviours are frequently associated in the literature with patients already diagnosed with AD; however, these changes are not always recognised as preceding diagnosis^46^. Regarding diet and nutrition, many other conditions, including type 1 diabetes, may exacerbate this effect, as we observed here up to 15 years before AD diagnosis^47^.

Changes in eating and drinking intake may also refer to changes in behaviours, therefore serving as an important potential screening tool or flag for early identification and intervention^48^. It remains unclear whether the coding of food and fluid intake symptoms refer to time during hospital admission or what contributed to hospital admission. Additional validation is required, with comparative datasets to explore this further.

### Interventional Implications

#### Timing

To ensure ongoing health and social care for populations living with dementia, it is essential to identify the factors that may contribute to their decline; hypothesis-driven findings, based on existing long-term conditions in populations, could modify disease progression and improve the outcomes. Similarly, the network analysis used here provides quantifiable measures (centrality) to help direct therapeutic management and prioritise a more tailored approach. Comorbidities including intestinal, circulatory and metabolic disorders associated with AD present earlier than the controls, matched for age and sex. However, VD comorbidity signatures are more persistent leading up to VD diagnosis, including circulatory, musculoskeletal, intestinal, respiratory and metabolic disorders that continue to present only in VD individuals. These conditions were also identified as having high central importance; therefore, prioritising management of these conditions in the appropriate time frame before or after dementia diagnosis could have a significant impact. With appropriate intervention and management of these conditions, clinicians may reduce downstream demand on healthcare services and redirect resources based on what has been observed in populations with AD and VD.

#### Long-term effects on healthcare

Considering downstream demand for healthcare services across AD and VD, patterns of mental health conditions were associated with each dementia sub-type. The VD cohort here showed higher variation in psychiatric conditions leading up to diagnosis, at closer time frames including depression and substance-related behavioural disorders such as those associated with alcohol and tobacco. Smoking is a common modifiable risk factor^8,49^. The extent to which this contributes to diagnosis of behavioural disorders resulting from tobacco use is less explored. Case control studies specifica lly analysing the effect of substance use but not behavioural disorder diagnosis on VD risk did not find similar associations^50^. Addressing substance use early would have wide implications for healthcare services and the health profile of populations at risk for dementia. Conditions such as depressive episodes were consistently characteristic of AD, earlier and longer than VD. Depressive symptoms have been associated with early markers of AD in comparative^51^ and case-control studies^52^. A meta-analysis found that a history of clinically associated depression may be associated with an increased risk for developing AD^53^ and using case-controlled cohorts^54^. More recent studies support the notion that greater depressive symptoms are associated with a higher incidence of dementia^8,55^. Longitudinal cohort studies suggest these symptoms may occur up to a decade before a diagnosis of dementia^56^, further supporting our findings in this study. Concurrent with depressive symptoms, apathy is a common neuropsychiatric symptom observed in people with dementia, often di”cult to disentangle with depressive symptoms^57^. A pilot study revealed that when compared with controls, dementia populations had significantly higher reports of apathy^58^. It is possible that coding used in hospital settings is not specific enough to distinguish these neuropsychiatric symptoms.

#### Immediate effects on healthcare

Complications and comorbidities, such as falls occurring close to dementia diagnosis, have a direct and immediate impact on health services and individuals’ quality of life^59^. In AD, there is a stronger association of delirium as an established intricate, interwoven relationship with one another^60^; those with a AD diagnosis have a higher risk of delirium, leading to a viscous cycle of worsening cognitive decline, further increasing risk^61^. It is suspected that a shared pathophysiological relationship exists between delirium and AD. The two conditions commonly co-occur, with correlations in blood biomarkers and inflammatory processes observed in both groups^62^.

The comorbid conditions identified in this work among dementia populations often serve as the basis for excluding participants from clinical and therapeutic trials developed for AD and VD^63^. If novel therapeutics are to be effective in the general population, it is important that representatives of characteristic pre-dementia pathologies are not excluded. Clinical trials that exclude people based on specific patterns of comorbidities may not accurately represent the heterogeneous AD population^64^, resulting in bias in trial results. Therefore, eligibility criteria for upcoming therapeutic targets in AD need to be more reflective of the population.

Post-diagnosis, we observed prominence of respiratory infections and complications for AD and VD. With greater management of these conditions, downstream effects on other complications including injury, falls and infections can be reduced or even mitigated^61,65^. Chronic respiratory infections, pneumonitis and skin ulcers and sores were identified as characteristic of AD and VD cohorts. The latter may be consequences of other conditions occurring earlier in disease trajectory. Our previous work has shown that over longitudinal observations of people living with dementia, those at a more advanced stage of dementia diagnosis rely heavily on community services^66^. This targeted identification of complications, symptoms, and conditions occurring after diagnosis of each dementia sub-type allows for a more focused approach to how community services can be resourced in dementia care to avoid the downstream effects of hospital admissions and the demand for primary care services.

### Limitations and Future Work

#### Dataset and population

Extracting accurate comorbidity diagnoses from this UK Biobank population poses a limitation for this work. The UK Biobank dataset provides three sources of data, in-patient hospital records, primary care and mortality data. The UK Biobank dataset reports the incidence of comorbidity diagnoses with a median delay of approximately 2.25 years; therefore, this may under-represent early-stage cases of each condition. However, it offers a suitable source to investigate temporal effects and impacts of comorbidities in dementia sub-types^67^. Hence, due to lack of complete data across all three data sources, we prioritised in-patient records to maximise sample size and capture conditions with the greatest impact on hospital services. Hospital admissions often mark critical inflection points where disease progression requires intervention, making them key for identifying uncontrolled or advanced disease. This approach focused on the most clinically significant events, enhancing the understanding of dementia-associated comorbidities and their healthcare demand.

As with all analyses based on electronic health records, this study is subject to known limitations related to the accuracy and completeness of diagnostic event timing. One such limitation involves so-called “dump” events, where a large number of diagnostic codes are entered on a single date. This often reflects the operational reality in clinical settings, where healthcare professionals may have limited, episodic interactions with the individual records. As a result, multiple pre-existing conditions may be retrospectively recorded during a single hospital admission or clinical encounter, leading to potential misclassification of disease onset. Furthermore, diagnostic information is frequently documented in alternative, unstructured formats such as clinical notes, particularly in multidisciplinary care settings. These free-text notes, which are not accessible via the UK Biobank dataset, likely contain clinically meaningful information that could enhance diagnostic accuracy and temporal resolution. Future research should explore the integration of such unstructured data, pending appropriate ethical approvals and governance frameworks.

The majority of this cohort had indicated their ethnicity as British (Figure 5). We did not integrate imputation for sampling bias as we wanted to reflect the current dementia population with in-patient records available. It has been shown that there is an inequality across the UK population regarding ethnic minorities. This includes the experience of minority groups in dementia care, from identification and diagnosis through to treatment, medications and hospitalisation^68–70^.

#### Technical limitations

In this analysis, we used using UK Biobank standard polygenic risk scores for the Alzheimer’s Disease trait and showed the polygenic risk scores for controls, compared with dementia sub-type groups^71,72^. However, future research should consider these risk scores in matching the disease and control groups at an individual level. This analysis did not compare replication with another population dataset; future research should aim to assess the reproducibility of results in other longitudinal cohorts. Analyses here did not directly confound for medications, and this would be an important next step in our work. Understanding what might be shifting towards a more ‘protective’ profile due to comorbidity diagnoses, prescribed medications, and management of comorbidities is important with regard to clinical interpretability. We conducted an undirected Bayesian network analysis. This approach does not consider the directional nature of conditions, such as how one may lead to another, exacerbate or protect. Therefore, this complex mapping remains to be further understood. Our aim in this work was not to determine the relative risk of each condition to either AD or VD. We applied methods to quantify relationships or associations. Although more basic, the Mann-Whitney U test offered a simpler starting point to establish a raw association. Building on this, logistic regression models confounded for factors such as sex and age at the time of dementia diagnosis. Finally, using network analysis, the wider context and accounting for different probabilities of conditions and how they relate to or influence one another were added. Collectively, this builds a comprehensive overview of the comorbidities that precede and follow AD and VD.

## Methods

### Study Design and Data Processing

This cohort study is based on data from the UK Biobank, a large-scale biomedical database containing genetic, lifestyle and health information from 446,848 participants, in the United Kingdom^73^, aged 40–73 years, who underwent assessment at one of 22 centres across the UK between 2006 and 2010. During the study, participants were asked to provide biological samples and complete questionnaires on an electronic tablet about a range of topics, including their demographics and underwent physical examination. Participants’ hospital inpatient records, coded by ICD-10 codes, were available as part of the UK Biobank dataset for 446,814 participants. These codes referred only to the first-instance diagnosis of each condition. This was in isolation from primary care records; we restricted the analysis to hospital inpatient records only as a large number of participants in the AD and VD cohort did not have primary care record data.

The UK Biobank electronic healthcare record data were collected following informed consent obtained from all participants. The North West Multi-centre Research Ethics Committee (MREC) and Research Tissue Bank (RTB) approved the scientific protocol and operational procedures (REC reference number: 16/NW/027) of the UK Biobank. Data for this study were obtained, and research was conducted under the UK Biobank applications license numbers (69610 and 109607).

### Covariates

Baseline characteristics extracted from the UK Biobank dataset included sex, year of birth, ethnicity and date of death (if applicable). Reasons for death were also available but were inconsistently recorded for our cohort and, therefore, not included in the analysis. The analysis was adjusted for age and sex, where appropriate.

### Cohort selection

Within the UK Biobank, there are pre-existing datasets that define a population. An example of this is the ‘All Cause Dementia’ dataset. However, this data included participants who self-reported their dementia diagnosis. We emphasised formal diagnosis and focused on in-patient records; therefore, we defined our cohort from ICD-10 coding instead.

### Dementia diagnosis

From the UK Biobank cohort, individuals had multiple diagnoses of dementia sub-types; therefore, we established a workflow to distinguish the most likely diagnosis. This process was similar to other work in the Scottish sub-set of the UK Biobank population, where coded diagnoses were compared across multiple primary care, inpatient and mortality data, with review by clinicians and dementia experts^74^.

We first defined a subset of ICD-10 codes to determine dementia diagnosis using ICD-10 classification, version 2019^75^: ‘Alzheimer’s Disease’: F00’, ‘F001’, ‘F002’, ‘F009’, ‘G30’, ‘G301’, ‘G308’, ‘G309’. ‘Vascular Dementia’: ‘F01’, ‘F011’, ‘F012’, F018’, ‘F019’. ‘Mixed Dementia’: ‘F013’. ‘Other’: ‘F02’, ‘F021’, ‘F022’, ‘F023’, ‘F024’, ‘F028’, ‘G31’. ‘Unspecified Dementia’: ‘F03’. ‘Mild Cognitive Impairment’: ‘F067’. We excluded the code ‘F05’ that was delirium-related conditions, not a dementia diagnosis. We also excluded the code ‘Q90’ corresponding to a diagnosis of ‘Down’s Syndrome’. This extracted 1O1,721 participants from the dataset. Participants who did not have in-patient hospital data available were also excluded, resulting in a final cohort of 9,834 potential participants. From the selected list of codes, we identified individuals with multiple dementia diagnoses in their records.

To accurately capture the clinical diagnosis of participants with more than one dementia diagnosis in their records, we established a standardised and reproducible approach, guided by a group of clinical experts within the Surrey and Borders Partnership NHS Foundation Trust. The clinical team comprised of Consultant Old Age Psychiatrists and dementia clinical experts mapped each participant to an ‘Overall Diagnosis’ from multiple ICD-10 codes. The criteria were as follows: Alzheimer’s Disease and Vascular Dementia were considered ‘primary’ diagnoses. Mixed Dementia, Mild Cognitive Impairment, and Unspecified Dementia were considered as ‘secondary’ diagnoses. All remaining diagnoses were collectively named ‘Other’, including Lewy Body Dementia, Dementia in Parkinson’s and Dementia in Huntington’s were considered ‘secondary’ diagnoses.

From the lists of multiple diagnosis per individual, we created a pipeline that established an ‘Overall Diagnosis’ for each participant, made up of: AD, VD, Mixed Dementia (made up of a mix of AD and VD), Mild Cognitive Impairment, Unspecified Dementia and Other. We excluded all diagnoses except AD and VD based on clinical uncertainty from data coding. Therefore, counts of each primary diagnosis were calculated for each individual, and the majority diagnosis was mapped as ‘Overall Diagnosis’ for each participant. For Mixed Dementia diagnoses, these were assigned to either AD or VD, depending on whether AD or VD count was higher. AD was assumed in all instances where: AD was the most prevalent diagnosis, AD was not present but Mixed Dementia count was more than VD, equal counts of either AD or VD and equal counts of AD and Mixed Dementia. VD was assumed if VD was the most prevalent diagnosis and if there were equal counts of VD and Mixed Dementia. Overall, this resulted in a cohort of participants who had a diagnosis of either Alzheimer’s Disease (n=3867) or Vascular Dementia (n=1481), a total cohort of 5,348 participants with AD or VD. A schematic of this process is detailed in Figure 6.

**Figure 6:**
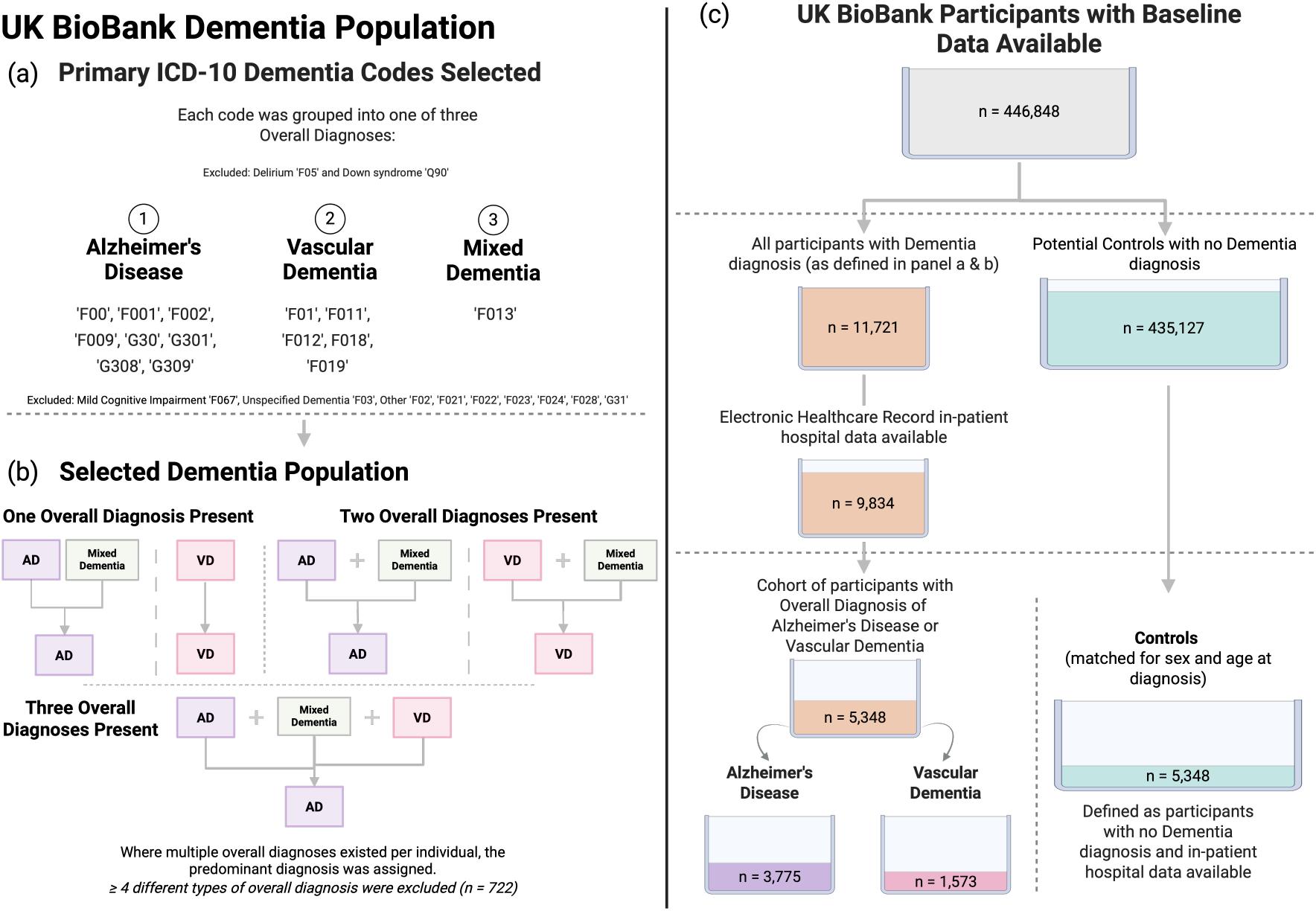
Schematic for the selection process of individuals from the UK Biobank dataset. (a) ICD-10 codes used to identify potential dementia diagnoses and how they were grouped into an ‘Overall Diagnosis’. (b) Participants with multiple dementia subtype codes were classified as either Alzheimer’s Disease or Vascular Dementia, according to the predominant diagnosis recorded. (c) Overview of data availability in the UK Biobank and the final cohort composition, including dementia subtypes and matched controls. (AD: Alzheimer’s Disease, VD: Vascular Dementia).

### Diagnosis date

The first date of each dementia diagnosis was used as the dementia onset date. This was also true for participants with multiple diagnoses in their records. This date was then assigned to the matched control as a ‘placeholder diagnosis flag’ date.

### Control Cohort

Once the cohort of participants with dementia was established (n=5,348), a systematic matching process was employed based on sex, year of birth, and date of death. This method incorporated a fixed random seed for random sampling and additional validation steps to account for age-specific effects while controlling for demographic and survival-related differences. Dementia participants were grouped based on their sex and age at diagnosis. These two variables were selected as primary matching criteria to control for demographic factors that could influence dementia risk or survival outcomes. Using UK Biobank standard polygenic risk scores for the Alzheimer’s Disease trait, we considered the polygenic risk scores for controls, compared with dementia sub-type groups^71,72^. We compared polygenic risk scores between groups using a two-sided Mann–Whitney U test due to non-normal distributions, and applied Benjamini–Hochberg correction to control the false discovery rate across multiple comparisons.

For potential controls, data from people with a diagnosis of dementia were removed from the main UK Biobank dataset, leaving 490,467 potential participants. Controls were defined as individuals of the same sex and age as the matched person with dementia who did not have a diagnosis of dementia in their records at that time. This ensured that the control group were an independent population from the dementia group. Additionally, any individuals with missing data across in-patient healthcare record data were removed.

Each dementia case had a known age at diagnosis that was derived from their year of birth and the year of dementia diagnosis. The age at diagnosis for controls was calculated using the year of diagnosis (dementia case) minus the year of birth, for control. Controls that had already been matched to another dementia case were excluded to ensure each control was used only once. We required that controls either had no recorded date of death or had a later date of death than the matched dementia case. As there were multiple potential controls available, we first selected up to three potential matched controls for each dementia individual and then used random selection to obtain a singular 1:1 ratio of matched controls from the set of three, using a fixed random seed to ensure reproducibility. Combined with matched controls, the final cohort for this analysis was 10,508 participants. Throughout the analysis, the same control case was assigned to the same dementia case to ensure a consistent comparison.

### Control disease

To separate dementia-specific patterns from general patterns of how comorbidities affect individuals as they age, a repeat analysis was conducted on a cohort of people diagnosed with hip fractures (ICD-10 code: ‘*S*72’. This was selected as a control disease due to its prevalence in older adults. It was also selected because it is a non-neurological condition with shared risk factors with dementia due to its association with ageing. Comparison across all dementia cohorts, controls and individuals with hip fractures, were all controlled for age at diagnosis and sex. For each individual with hip fracture, we matched them with an individual without a diagnosis of hip fracture, as well as, excluding all individuals already used in dementia and control cohorts previously.

### Categorising Comorbidities Using ICD-10 Codes, Blocks and Chapters

The ICD-10 coding system is a hierarchical system for categorising diseases, comorbidities and complications of disease. The top level are Chapters (1-22), followed by blocks (A00-Z99; approximately 221), which further classify diseases within each Chapter. Following blocks, there are Categories and Sub-categories. The UK Biobank dataset provides the individual ICD-10 codes for the first instance of each diagnosis per participant; these individual codes were the primary source for this analysis. Counts of each individual diagnosis were calculated for dementia and control groups. We further categorised each individual code into its corresponding block and chapter using the World Health Organisation ICD-10 Application Programme Interface (API), 2019 release^75^.

### Chronological mapping of comorbidities

Using ‘days to dementia diagnosis’, all ICD-10 diagnoses of comorbidities were mapped relative to the central dementia diagnosis date. For controls, a ‘placeholder diagnosis flag’ date was assigned based on their matched dementia participant. Mappings were grouped into categorical time frames. For those before dementia: 20+ years before, 15-20 years before, 10-15 years before, 7-10 years before, 5-7 years before, 2-5 years before, and 0-2 years before, reducing the time frame span, as it got closer to the diagnosis date. For after dementia diagnosis, these were categorised into four time-frames: 0-2 years after, 0-4 years after, 0-6 years after, 0-8 years after and 0-10 years after. We identified a condition as ‘new’ if it had not already been diagnosed in previous years. Comorbidities have an important cumulative effect over time, and thus, it is not suitable to consider conditions only according to the time frame in which they first occur. They must also be considered over the time following emergence and in relation to each other. An assumption was made, such that from diagnosis of each comorbidity, that diagnosis remained with that individual. If there was a repeat diagnosis, this was counted in addition to the previous diagnosis. It is worth noting that clinically, not all comorbidities are alike; some are diagnosed, managed and resolved and some are unresolved or become chronic. However, for the purposes of this analysis, all conditions were considered to remain present following initial diagnosis and were not removed from individual records. This will reduce the risk of underestimating the effect of comorbidities. This approach also reflects the reality of conditions, especially in older populations, which are often progressive, chronic or complex with long-lasting effects.

### Statistical Analysis & Machine Learning

To assess the differences in prevalence of comorbidities between groups, we used the two-sided Mann-Whitney U Test^76^, a non-parametric test to compare two independent samples. We applied the Bonferroni correction^77^ to control for Type I errors. Firstly, this analysis was conducted using individual diagnoses of ICD-10 conditions, using a binary system for presence or absence of the condition for each individual. A second analysis was conducted in parallel, using the sum of each ICD-10 block per individual.

### Regression analyses of Alzheimer’s Disease and Vascular Dementia

A logistic regression analysis was performed for each time frame, separating models for AD and VD. This was only conducted for time frames prior to diagnosis. We included both age at diagnosis and age at diagnosis (squared) to assess potential non-linear effects of age on risk of dementia sub-type diagnosis. We also included an interaction term (age at diagnosis and sex) to evaluate potential age-at-diagnosis-by-sex interaction effects. The performance of each model was determined using the Area Under the Curve (AUC). A range of different models were built to establish the best performing, including logistic regression with *L*1 regularisation, logistic regression with *L*2 regularisation, Random Forest and XGBoost. Odds ratios and corresponding confidence intervals for each condition and its association with AD or VD were calculated. Conditions were determined to be significant if the *p-value* < 0.05.

### Analysis of comorbidity networks across time points

To account for the impact of comorbidities across an individual’s health landscape leading up to dementia diagnosis, we conducted an undirected network analysis^78^. The network analysis was performed for each time frame and each dementia sub-type. Undirected graphical models are also known as Markov Random Fields. Markov Random Fields are similar to Bayesian networks but differ in that they use undirected edges to represent non-causal dependencies. These probabilistic models can represent correlations and conditional dependencies between variables in complex systems. In the network analysis, and for each time frame, at least 100 different individuals were included for both dementia and control cohorts. This was conducted using only the conditions most significantly (*p*-value <0.05) associated with a dementia diagnosis, as determined by the two-sided Mann-Whitney U test.

A minimum support threshold of 50 was used for ICD-10 individual diagnostic codes. However, this was increased to 100 for analysis of ICD-10 blocks. These thresholds were chosen to ensure the analysis focused on clinically insightful patterns while reducing noise from infrequent combinations. Centrality measures were calculated for the conditions (i.e. nodes) within each network, specific to time frame and dementia sub-type, and replicated for matched controls. Each diagnosis, per individual, was categorised into a time frame relative to the date of first diagnosis of dementia and split to AD or VD. The matched controls were assigned to the dementia sub-type in which their corresponding dementia participant was. Once the networks for each time frame and subtype were established, the networks were compared across the time frames. Conditions unique to the first network (20+ years before diagnosis) were identified across the dementia and control groups and marked as re-occurring when identified across consecutive time frames. Therefore, newly diagnosed conditions were identified at each time frame per dementia sub-type. The same analysis was repeated for ICD-10 blocks, with a minimum support of 100.

### Time to death analysis

Survival analysis was conducted, and Hazard Ratio (HR)s were calculated using the Cox Proportional Hazards regression model^79^ with time to death as the dependent variable. The analysis incorporated covariates of age and sex, as well as the different dementia subtypes according to ‘Overall Diagnosis’, as described above. HR for each of the ‘Overall Diagnosis’ groups were calculated, and the time to death at survival probability intervals was calculated for the control cohort. For AD and VD, the time in months that the dementia cohort was ahead of the controls was calculated; i.e. an individual with dementia would die *X* months sooner than a control individual of the same sex and age at the time of the dementia diagnosis.

## Data Availability

The data that support the findings of this study are available from the UK Biobank, found here: https://www.ukbiobank.ac.uk. The cohort populations that we have identified will be also made available from UK Biobank, upon publication.

The code used in this study will be made available by the corresponding author on reasonable request.

## Acknowledgements

This research has been conducted using the UK Biobank Resource under Application Numbers 69610 and 109607. We would like to acknowledge support from Surrey and Borders Partnership NHS Foundation Trust and the UK Dementia Research Institute, Care Research and Technology Centre.

This study is funded by the UK Dementia Research Institute (UK DRI) Care Research and Technology Centre funded by the Medical Research Council (MRC), Alzheimer’s Research UK, Alzheimer’s Society (grant number: UKDRI–7002), and the UKRI Engineering and Physical Sciences Research Council (EPSRC) Resilient Project (grant number: EP/W031892/1). Infrastructure support for this research was provided by the NIHR Imperial Biomedical Research Centre (BRC) and the UKRI Medical Research Council (MRC). P.B. is funded by Great Ormond Street Hospital and the Royal Academy of Engineering.

C.S. and A.K.S are supported by the UK Dementia Research Institute [UK DRI-5209] and a UKRI Future Leaders Fellowship [MR/X032892/1]. C.S. also receives personal support from the Edmond J. Safra Foundation.

## Author Contributions

**CW:** Conceptualisation, Methodology, Software, Formal Analysis, Investigation, Data Curation, Validation, Visualisation, Writing - Original Draft, Review and Editing; **AF** Software, Review and Editing; **AKS** Software, Review and Editing; **VM** Writing - Original Draft, Review and Editing; **BS** Writing - Original Draft, Review and Editing; **CS** Review and Editing; **MR** Review and Editing; **RN:** Conceptualisation, Methodology, Writing - Original Draft, Review and Editing, Supervision; **PB:** Conceptualisation, Methodology, Writing - Original Draft, Review and Editing, Supervision, Funding Acquisition.

## Abbreviations

AD: Alzheimer’s Disease
HR: Hazard Ratio
ICD-10: International Classification of Disease - 10
NHS: The UK National Health Service
OR: Odds Ratio
VD: Vascular Dementia

