## Supplementary Information for "Chronological Mapping of Comorbidities in Alzheimer’s Disease and Vascular Dementia"

#### Supplementary Results

Several models were used to establish the predictive ability of conditions diagnosed in each time frame, separately for AD and VD (Supplementary Table S1 and S2). Logistic regression with L1 regularisation was selected as the most appropriate model. This was the best performing model for VD overall, and there were small differences between this model and other models for AUC in the AD cohort. In addition, VD cohorts have shown more distinct patterns of conditions over time, particularly leading up to diagnosis. Therefore, as we did not aim to predict if people will be diagnosed with either AD or VD, we only set out to determine the predictive ability of conditions with respect to dementia sub-type; performance was not prioritised for model selection.

#### Supplementary Tables

**Table S1: Model performance metrics for classification machine learning models for Alzheimer's Disease.** Comorbidity diagnosis as inputs, confounding for age at diagnosis of Alzheimer's Disease and sex. (AD: Alzheimer's Disease, AUC: Area under Curve).

| Time Frame | Model Type | Sensitivity | Specificity | Accuracy | Precision | F1-Score | AUC |
|---|---|---|---|---|---|---|---|
| 20+ years before AD | Logistic Regression (L1) | 0.407 | 0.605 | 0.503 | 0.521 | 0.457 | 0.505 |
| 20+ years before AD | Logistic Regression (L2) | 0.824 | 0.174 | 0.508 | 0.514 | 0.633 | 0.491 |
| 20+ years before AD | Random Forest | 0.505 | 0.535 | 0.520 | 0.535 | 0.520 | 0.526 |
| 20+ years before AD | XGBoost | 0.505 | 0.512 | 0.508 | 0.523 | 0.514 | 0.498 |
| 20-15 years before AD | Logistic Regression (L1) | 0.563 | 0.447 | 0.512 | 0.568 | 0.565 | 0.522 |
| 20-15 years before AD | Logistic Regression (L2) | 0.918 | 0.0894 | 0.556 | 0.565 | 0.700 | 0.496 |
| 20-15 years before AD | Random Forest | 0.558 | 0.475 | 0.522 | 0.578 | 0.568 | 0.543 |
| 20-15 years before AD | XGBoost | 0.524 | 0.536 | 0.529 | 0.593 | 0.556 | 0.531 |
| 15-10 years before AD | Logistic Regression (L1) | 0.575 | 0.439 | 0.511 | 0.534 | 0.554 | 0.505 |
| 15-10 years before AD | Logistic Regression (L2) | 0.808 | 0.190 | 0.516 | 0.527 | 0.638 | 0.494 |
| 15-10 years before AD | Random Forest | 0.541 | 0.480 | 0.512 | 0.537 | 0.539 | 0.478 |
| 15-10 years before AD | XGBoost | 0.536 | 0.472 | 0.506 | 0.531 | 0.534 | 0.509 |
| 10-7 years before AD | Logistic Regression (L1) | 0.549 | 0.540 | 0.544 | 0.538 | 0.543 | 0.560 |
| 10-7 years before AD | Logistic Regression (L2) | 1.000 | 0.000 | 0.494 | 0.494 | 0.661 | 0.528 |
| 10-7 years before AD | Random Forest | 0.522 | 0.481 | 0.501 | 0.495 | 0.508 | 0.516 |
| 10-7 years before AD | XGBoost | 0.511 | 0.544 | 0.528 | 0.522 | 0.517 | 0.530 |
| 7-5 years before AD | Logistic Regression (L1) | 0.438 | 0.677 | 0.548 | 0.611 | 0.510 | 0.578 |
| 7-5 years before AD | Logistic Regression (L2) | 0.531 | 0.525 | 0.528 | 0.565 | 0.548 | 0.534 |
| 7-5 years before AD | Random Forest | 0.538 | 0.540 | 0.539 | 0.576 | 0.557 | 0.557 |
| 7-5 years before AD | XGBoost | 0.411 | 0.708 | 0.548 | 0.621 | 0.494 | 0.579 |
| 5-2 years before AD | Logistic Regression (L1) | 0.486 | 0.707 | 0.596 | 0.629 | 0.549 | 0.622 |
| 5-2 years before AD | Logistic Regression (L2) | 0.435 | 0.683 | 0.558 | 0.583 | 0.499 | 0.582 |
| 5-2 years before AD | Random Forest | 0.577 | 0.600 | 0.589 | 0.595 | 0.586 | 0.612 |
| 5-2 years before AD | XGBoost | 0.472 | 0.730 | 0.600 | 0.641 | 0.544 | 0.639 |
| 2 years before AD | Logistic Regression (L1) | 0.472 | 0.756 | 0.614 | 0.657 | 0.549 | 0.639 |
| 2 years before AD | Logistic Regression (L2) | 0.534 | 0.473 | 0.504 | 0.502 | 0.518 | 0.509 |
| 2 years before AD | Random Forest | 0.533 | 0.675 | 0.604 | 0.620 | 0.573 | 0.644 |
| 2 years before AD | XGBoost | 0.421 | 0.807 | 0.615 | 0.685 | 0.522 | 0.651 |

**Table S2: Model performance metrics for classification machine learning models for Vascular Dementia.** Comorbidity diagnosis as inputs, confounding for age at diagnosis of Vascular Dementia and sex. (VD: Vascular Dementia, AUC: Area under Curve).

| Time Frame | Model Type | Sensitivity | Specificity | Accuracy | Precision | F1-Score | AUC |
|---|---|---|---|---|---|---|---|
| 20+ years before VD | Logistic Regression (L1) | 0.907 | 0.091 | 0.671 | 0.710 | 0.797 | 0.621 |
| 20+ years before VD | Logistic Regression (L2) | 1.000 | 0.000 | 0.711 | 0.711 | 0.831 | 0.499 |
| 20+ years before VD | Random Forest | 0.704 | 0.182 | 0.553 | 0.679 | 0.691 | 0.487 |
| 20+ years before VD | XGBoost | 0.778 | 0.182 | 0.605 | 0.700 | 0.737 | 0.526 |
| 20-15 years before VD | Logistic Regression (L1) | 0.853 | 0.229 | 0.599 | 0.617 | 0.716 | 0.559 |
| 20-15 years before VD | Logistic Regression (L2) | 1.000 | 0.000 | 0.593 | 0.593 | 0.745 | 0.540 |
| 20-15 years before VD | Random Forest | 0.755 | 0.371 | 0.599 | 0.636 | 0.691 | 0.571 |
| 20-15 years before AD | XGBoost | 0.745 | 0.300 | 0.564 | 0.608 | 0.670 | 0.583 |
| 15-10 years before VD | Logistic Regression (L1) | 0.695 | 0.482 | 0.604 | 0.644 | 0.668 | 0.660 |
| 15-10 years before VD | Logistic Regression (L2) | 1.000 | 0.000 | 0.574 | 0.574 | 0.729 | 0.515 |
| 15-10 years before VD | Random Forest | 0.721 | 0.539 | 0.644 | 0.678 | 0.699 | 0.646 |
| 15-10 years before VD | XGBoost | 0.689 | 0.567 | 0.637 | 0.682 | 0.686 | 0.658 |
| 10-7 years before VD | Logistic Regression (L1) | 0.638 | 0.583 | 0.614 | 0.661 | 0.649 | 0.648 |
| 10-7 years before VD | Logistic Regression (L2) | 0.659 | 0.522 | 0.599 | 0.637 | 0.648 | 0.644 |
| 10-7 years before VD | Random Forest | 0.603 | 0.583 | 0.594 | 0.648 | 0.624 | 0.648 |
| 10-7 years before VD | XGBoost | 0.620 | 0.578 | 0.601 | 0.651 | 0.635 | 0.636 |
| 7-5 years before VD | Logistic Regression (L1) | 0.594 | 0.668 | 0.626 | 0.704 | 0.644 | 0.679 |
| 7-5 years before VD | Logistic Regression (L2) | 0.855 | 0.233 | 0.588 | 0.597 | 0.703 | 0.511 |
| 7-5 years before VD | Random Forest | 0.688 | 0.560 | 0.633 | 0.674 | 0.681 | 0.703 |
| 7-5 years before VD | XGBoost | 0.598 | 0.668 | 0.628 | 0.705 | 0.647 | 0.690 |
| 5-2 years before VD | Logistic Regression (L1) | 0.629 | 0.771 | 0.692 | 0.775 | 0.694 | 0.737 |
| 5-2 years before VD | Logistic Regression (L2) | 0.575 | 0.722 | 0.640 | 0.722 | 0.640 | 0.699 |
| 5-2 years before VD | Random Forest | 0.686 | 0.700 | 0.692 | 0.741 | 0.712 | 0.725 |
| 5-2 years before VD | XGBoost | 0.614 | 0.803 | 0.698 | 0.796 | 0.694 | 0.754 |
| 2 years before VD | Logistic Regression (L1) | 0.708 | 0.829 | 0.766 | 0.815 | 0.758 | 0.821 |
| 2 years before VD | Logistic Regression (L2) | 0.657 | 0.755 | 0.704 | 0.741 | 0.696 | 0.773 |
| 2 years before VD | Random Forest | 0.719 | 0.770 | 0.744 | 0.770 | 0.743 | 0.816 |
| 2 years before VD | XGBoost | 0.690 | 0.856 | 0.770 | 0.836 | 0.756 | 0.814 |

**Table S3: Significantly associated comorbidities with Alzheimer's Disease cohort from logistic regression model.** Significance here refers to *p-value* <0.05. Conditions are displayed with corresponding odds ratios and 95% confidence intervals. Conditions are displayed in alphabetical order.

| Time Frame | Condition | Odds Ratio | CI Lower | CI Upper | *p-value* |
|---|---|---|---|---|---|
| 20+ years before | Ileus | 0.216 | 0.072 | 0.651 | 0.007 |
| 20-15 years before | Bacterial intestinal infection | 4.243 | 1.071 | 16.808 | 0.040 |
| 20-15 years before | Chest pain | 1.517 | 1.067 | 2.157 | 0.020 |
| 20-15 years before | Generalized and unspecified atherosclerosis | 3.283 | 1.145 | 9.414 | 0.027 |
| 15-10 years before | Abnormal finding of blood chemistry | 0.505 | 0.285 | 0.893 | 0.019 |
| 15-10 years before | Oedema | 0.187 | 0.048 | 0.732 | 0.016 |
| 15-10 years before | Personal history of other specified conditions | 0.717 | 0.534 | 0.961 | 0.026 |
| 15-10 years before | Problem related to lifestyle | 1.553 | 1.048 | 2.300 | 0.028 |
| 15-10 years before | Unknown and unspecified causes of morbidity | 1.369 | 1.003 | 1.867 | 0.048 |
| 15-10 years before | Unspecified haematuria | 0.652 | 0.473 | 0.899 | 0.009 |
| 15-10 years before | Unspecified visual impairment (binocular) | 5.064 | 1.421 | 18.048 | 0.012 |
| 10-7 years before | Abnormal finding of blood chemistry | 0.587 | 0.385 | 0.894 | 0.013 |
| 10-7 years before | Arthrosis | 1.366 | 1.063 | 1.756 | 0.015 |
| 10-7 years before | Chest pain | 1.258 | 1.039 | 1.522 | 0.019 |
| 10-7 years before | Depressive episode | 1.599 | 1.047 | 2.441 | 0.030 |
| 10-7 years before | Extrapyramidal and movement disorders | 4.537 | 1.282 | 16.051 | 0.019 |
| 10-7 years before | Intervertebral disc disorder | 1.590 | 1.010 | 2.501 | 0.045 |
| 10-7 years before | Other and unspecified symptoms and signs involving cognitive functions and awareness | 2.536 | 1.122 | 5.730 | 0.025 |
| 10-7 years before | Other symptoms and signs concerning food and fluid intake | 2.610 | 1.641 | 4.152 | 0.000 |
| 10-7 years before | Problem related to lifestyle | 1.536 | 1.148 | 2.055 | 0.004 |
| 10-7 years before | Special screening examination | 1.589 | 1.044 | 2.420 | 0.031 |
| 10-7 years before | Type 2 diabetes mellitus without complications | 1.312 | 1.024 | 1.681 | 0.032 |
| 7-5 years before | Degenerative disease of nervous system | 6.905 | 1.872 | 25.476 | 0.004 |
| 7-5 years before | Depressive episode | 1.847 | 1.292 | 2.642 | 0.001 |
| 7-5 years before | Extrapyramidal and movement disorders | 3.053 | 1.130 | 8.250 | 0.028 |
| 7-5 years before | Oedema | 0.241 | 0.088 | 0.658 | 0.006 |
| 7-5 years before | Other and unspecified speech disturbances | 0.263 | 0.101 | 0.685 | 0.006 |
| 7-5 years before | Other and unspecified symptoms and signs involving cognitive functions and awareness | 2.973 | 1.666 | 5.306 | 0.000 |
| 7-5 years before | Other symptoms and signs concerning food and fluid intake | 1.905 | 1.305 | 2.782 | 0.001 |
| 7-5 years before | Problem related to lifestyle | 1.425 | 1.079 | 1.882 | 0.013 |
| 7-5 years before | Unknown and unspecified causes of morbidity | 1.352 | 1.013 | 1.805 | 0.040 |
| 7-5 years before | Unspecified haematuria | 0.759 | 0.585 | 0.985 | 0.038 |
| 5-2 years before | Accidental poisoning by and exposure to other and unspecified drugs, medicaments and biological substances | 4.555 | 1.118 | 18.549 | 0.034 |
| 5-2 years before | Arthrosis | 1.195 | 1.007 | 1.417 | 0.041 |
| 5-2 years before | Chest pain | 1.360 | 1.160 | 1.594 | 0.000 |
| 5-2 years before | Degenerative disease of nervous system | 14.674 | 6.855 | 31.413 | 0.000 |
| 5-2 years before | Delirium | 4.103 | 1.775 | 9.483 | 0.001 |
| 5-2 years before | Depressive episode | 1.617 | 1.201 | 2.176 | 0.002 |
| 5-2 years before | Extrapyramidal and movement disorders | 5.210 | 2.299 | 11.808 | 0.000 |
| 5-2 years before | Functional intestinal disorder | 1.352 | 1.026 | 1.780 | 0.032 |
| 5-2 years before | Medical care | 0.771 | 0.623 | 0.954 | 0.017 |
| 5-2 years before | Oedema | 0.467 | 0.229 | 0.950 | 0.036 |
| 5-2 years before | Other and unspecified symptoms and signs involving cognitive functions and awareness | 4.716 | 3.122 | 7.125 | 0.000 |
| 5-2 years before | Other fall on same level | 2.043 | 1.105 | 3.774 | 0.023 |
| 5-2 years before | Other symptoms and signs concerning food and fluid intake | 2.067 | 1.547 | 2.760 | 0.000 |
| 5-2 years before | Pulmonary embolism without mention of acute cor pulmonale | 0.507 | 0.273 | 0.942 | 0.032 |
| 5-2 years before | Spondylopathy | 1.479 | 1.029 | 2.127 | 0.034 |
| 5-2 years before | Sprain and strain of other and unspecified parts of shoulder girdle | 5.773 | 1.824 | 18.269 | 0.003 |
| 5-2 years before | Unspecified haematuria | 0.778 | 0.607 | 0.997 | 0.047 |
| 2 years before to diagnosis | Bipolar affective disorder | 2.758 | 1.162 | 6.550 | 0.021 |
| 2 years before to diagnosis | Chest pain | 1.269 | 1.084 | 1.486 | 0.003 |
| 2 years before to diagnosis | Degenerative disease of nervous system | 30.307 | 15.258 | 60.198 | 0.000 |
| 2 years before to diagnosis | Delirium | 4.945 | 3.033 | 8.062 | 0.000 |
| 2 years before to diagnosis | Depressive episode | 1.525 | 1.169 | 1.988 | 0.002 |
| 2 years before to diagnosis | Extrapyramidal and movement disorders | 3.898 | 2.079 | 7.309 | 0.000 |
| 2 years before to diagnosis | Oedema | 0.471 | 0.244 | 0.909 | 0.025 |
| 2 years before to diagnosis | Other and unspecified symptoms and signs involving cognitive functions and awareness | 6.788 | 4.734 | 9.732 | 0.000 |
| 2 years before to diagnosis | Other and unspecified symptoms and signs involving the nervous and musculoskeletal systems | 1.472 | 1.033 | 2.096 | 0.032 |
| 2 years before to diagnosis | Other fall on same level | 1.648 | 1.003 | 2.708 | 0.048 |
| 2 years before to diagnosis | Other symptoms and signs concerning food and fluid intake | 2.294 | 1.741 | 3.021 | 0.000 |
| 2 years before to diagnosis | Personal history of allergy to unspecified drugs, medicaments and biological substances | 0.851 | 0.747 | 0.970 | 0.015 |
| 2 years before to diagnosis | Procedure not carried out reason | 1.258 | 1.057 | 1.499 | 0.010 |
| 2 years before to diagnosis | Special screening examination | 1.354 | 1.047 | 1.752 | 0.021 |
| 2 years before to diagnosis | Unknown and unspecified causes of morbidity | 1.475 | 1.144 | 1.901 | 0.003 |
| 2 years before to diagnosis | Unspecified fall | 1.578 | 1.111 | 2.243 | 0.011 |

**Table S4: Significantly associated comorbidities with Vascular Dementia cohort from logistic regression model.** Significance here refers to $p$-*value* <0.05. Conditions are displayed with corresponding odds ratios and 95% confidence intervals. Conditions are displayed in alphabetical order.

| Time Frame | Condition | Odds Ratio | CI Lower | CI Upper | $p$-*value* |
|---|---|---|---|---|---|
| 20+ years before | Problem related to lifestyle | 3.697 | 1.130 | 12.097 | 0.031 |
| 20-15 years before | Care involving use of rehabilitation procedure | 4.458 | 1.002 | 19.839 | 0.050 |
| 20-15 years before | Cerebral infarction | 5.013 | 1.642 | 15.300 | 0.005 |
| 20-15 years before | Disorder of urinary system | 2.346 | 1.003 | 5.488 | 0.049 |
| 20-15 years before | Personal history of other specified conditions | 0.312 | 0.122 | 0.801 | 0.015 |
| 20-15 years before | Presence of cardiac and vascular implant and graft | 2.540 | 1.015 | 6.357 | 0.046 |
| 20-15 years before | Special screening examination | 3.641 | 1.075 | 12.327 | 0.038 |
| 20-15 years before | Type 2 diabetes mellitus without complications | 2.163 | 1.011 | 4.628 | 0.047 |
| 20-15 years before | Unknown and unspecified causes of morbidity | 2.043 | 1.043 | 4.002 | 0.037 |
| 15-10 years before | Abnormal finding of blood chemistry | 0.254 | 0.087 | 0.738 | 0.012 |
| 15-10 years before | Cerebral infarction | 2.691 | 1.064 | 6.803 | 0.036 |
| 15-10 years before | Cerebrovascular disease | 13.897 | 3.449 | 56.003 | 0.000 |
| 15-10 years before | Epilepsy | 4.197 | 1.533 | 11.494 | 0.005 |
| 15-10 years before | Personal history of diseases of the circulatory system | 1.588 | 1.131 | 2.229 | 0.008 |
| 15-10 years before | Rheumatoid arthritis | 4.307 | 1.605 | 11.559 | 0.004 |
| 15-10 years before | Type 2 diabetes mellitus without complications | 1.936 | 1.229 | 3.051 | 0.004 |
| 10-7 years before | Cerebral infarction | 3.891 | 1.562 | 9.695 | 0.004 |
| 10-7 years before | Cerebrovascular disease | 3.919 | 1.117 | 13.750 | 0.033 |
| 10-7 years before | Chest pain | 1.386 | 1.002 | 1.917 | 0.049 |
| 10-7 years before | Degenerative disease of nervous system | 4.006 | 1.007 | 15.933 | 0.049 |
| 10-7 years before | Dorsalgia | 1.576 | 1.035 | 2.400 | 0.034 |
| 10-7 years before | Epilepsy | 3.438 | 1.351 | 8.750 | 0.010 |
| 10-7 years before | Faecal incontinence | 4.489 | 1.040 | 19.368 | 0.044 |
| 10-7 years before | Fracture of foot | 3.810 | 1.004 | 14.453 | 0.049 |
| 10-7 years before | Hypertensive diseases | 1.330 | 1.030 | 1.717 | 0.029 |
| 10-7 years before | Hypotension | 3.010 | 1.180 | 7.677 | 0.021 |
| 10-7 years before | Personal history of diseases of the circulatory system | 1.399 | 1.080 | 1.813 | 0.011 |
| 10-7 years before | Type 2 diabetes mellitus without complications | 1.860 | 1.315 | 2.630 | 0.000 |
| 7-5 years before | Cerebral infarction | 3.600 | 1.522 | 8.518 | 0.004 |
| 7-5 years before | Cerebrovascular disease | 4.278 | 1.338 | 13.673 | 0.014 |
| 7-5 years before | Dorsalgia | 1.483 | 1.039 | 2.118 | 0.030 |
| 7-5 years before | Hypertensive diseases | 1.358 | 1.057 | 1.744 | 0.017 |
| 7-5 years before | Intracerebral haemorrhage | 8.412 | 3.406 | 20.779 | 0.000 |
| 7-5 years before | Medical care | 0.624 | 0.403 | 0.966 | 0.035 |
| 7-5 years before | Special screening examination | 2.086 | 1.102 | 3.949 | 0.024 |
| 7-5 years before | Sprain and strain of other and unspecified parts of shoulder girdle | 5.289 | 1.014 | 27.590 | 0.048 |
| 7-5 years before | Type 2 diabetes mellitus without complications | 1.520 | 1.093 | 2.114 | 0.013 |
| 5-2 years before | Bipolar affective disorder | 3.198 | 1.323 | 7.729 | 0.010 |
| 5-2 years before | Cerebral infarction | 3.163 | 1.624 | 6.160 | 0.001 |
| 5-2 years before | Cerebrovascular disease | 5.069 | 2.059 | 12.478 | 0.000 |
| 5-2 years before | Degenerative disease of nervous system | 3.843 | 1.251 | 11.803 | 0.019 |
| 5-2 years before | Dorsalgia | 1.521 | 1.042 | 2.220 | 0.030 |
| 5-2 years before | Hypertensive diseases | 1.489 | 1.156 | 1.919 | 0.002 |
| 5-2 years before | Intracerebral haemorrhage | 8.712 | 3.235 | 23.462 | 0.000 |
| 5-2 years before | Medical care | 0.614 | 0.420 | 0.898 | 0.012 |
| 5-2 years before | Mental and behavioural disorders due to use of tobacco : unspecified mental and behavioural disorder | 1.830 | 1.059 | 3.162 | 0.030 |
| 5-2 years before | Other and unspecified symptoms and signs involving cognitive functions and awareness | 3.365 | 1.643 | 6.892 | 0.001 |
| 5-2 years before | Schizophrenia | 5.901 | 1.634 | 21.302 | 0.007 |
| 5-2 years before | Special screening examination | 2.083 | 1.181 | 3.673 | 0.011 |
| 5-2 years before | Type 2 diabetes mellitus without complications | 2.105 | 1.519 | 2.918 | 0.000 |
| 5-2 years before | Unknown and unspecified causes of morbidity | 1.736 | 1.093 | 2.757 | 0.019 |
| 5-2 years before | Unspecified injury of head | 7.415 | 1.674 | 32.844 | 0.008 |
| 2 years before to diagnosis | Bipolar affective disorder | 3.141 | 1.204 | 8.195 | 0.019 |
| 2 years before to diagnosis | Cerebral infarction | 2.485 | 1.381 | 4.471 | 0.002 |
| 2 years before to diagnosis | Cerebrovascular disease | 3.032 | 1.529 | 6.012 | 0.001 |
| 2 years before to diagnosis | Degenerative disease of nervous system | 10.152 | 3.733 | 27.603 | 0.000 |
| 2 years before to diagnosis | Delirium | 6.368 | 2.490 | 16.285 | 0.000 |
| 2 years before to diagnosis | Depressive episode | 1.868 | 1.138 | 3.065 | 0.013 |
| 2 years before to diagnosis | Intracerebral haemorrhage | 20.171 | 8.659 | 46.985 | 0.000 |
| 2 years before to diagnosis | Medical care | 0.632 | 0.423 | 0.944 | 0.025 |
| 2 years before to diagnosis | Mental and behavioural disorders due to use of tobacco : unspecified mental and behavioural disorder | 1.967 | 1.101 | 3.515 | 0.022 |
| 2 years before to diagnosis | Other and unspecified symptoms and signs involving cognitive functions and awareness | 6.635 | 3.745 | 11.755 | 0.000 |
| 2 years before to diagnosis | Type 1 diabetes mellitus without complications | 2.435 | 1.019 | 5.818 | 0.045 |
| 2 years before to diagnosis | Type 2 diabetes mellitus without complications | 1.657 | 1.210 | 2.270 | 0.002 |
| 2 years before to diagnosis | Unknown and unspecified causes of morbidity | 1.899 | 1.168 | 3.087 | 0.010 |
| 2 years before to diagnosis | Unspecified fall | 1.870 | 1.042 | 3.356 | 0.036 |

**Table S5: Ethnicity data for dementia and control cohorts according to their dementia diagnosis type.** The control cohort's overall diagnosis here corresponds to their age- and sex-matched dementia participant.

| Overall Diagnosis | Ethnicity | Dementia Cohort Count | Control Cohort Count |
|---|---|---|---|
| Alzheimer's Disease | British | 3502 | 3527 |
| | Irish | 111 | 91 |
| | Any other white background | 84 | 98 |
| | Caribbean | 38 | 21 |
| | Indian | 26 | 37 |
| | Other ethnic group | 19 | 17 |
| | Prefer not to answer | 13 | 10 |
| | African | 12 | 14 |
| | Any other Asian background | 12 | 11 |
| | Missing | 11 | 7 |
| | White | 9 | 5 |
| | Pakistani | 8 | 5 |
| | Bangladeshi | 5 | 1 |
| | Any other mixed background | 4 | 1 |
| | Chinese | 4 | 9 |
| | White and Asian | 2 | 6 |
| | Do not know | 1 | 2 |
| | White and Black African | 1 | 5 |
| Vascular Dementia | British | 1327 | 1339 |
| | Irish | 55 | 35 |
| | Any other white background | 27 | 37 |
| | Caribbean | 16 | 8 |
| | Indian | 15 | 13 |
| | African | 11 | 7 |
| | Other ethnic group | 9 | 10 |
| | Missing | 6 | 3 |
| | Any other mixed background | 3 | 1 |
| | White | 3 | 2 |
| | Any other Asian background | 1 | 7 |
| | Pakistani | 1 | 6 |
| | Prefer not to answer | 1 | 5 |
| | White and Asian | 1 | 2 |
| | White and Black African | 1 | 1 |
| | White and Black Caribbean | 1 | 1 |

**Table S6: Kaplan-Meier Survival analysis for Alzheimer's Disease and Vascular Dementia cohort vs. controls.** Lag refers to how much earlier (in months) the dementia sub-type cohorts reached corresponding survival probabilities compared to controls. All controls were matched based on sex and age at diagnosis of dementia. All survival probabilities included. (AD: Alzheimer's Disease, VD: Vascular Dementia).

| Survival Probability | AD Lag (Months) | VD Lag (Months) |
|---|---|---|
| 0.10 | 25.63 | 32.97 |
| 0.20 | 14.93 | 23.90 |
| 0.30 | 15.57 | 24.50 |
| 0.40 | 14.73 | 22.20 |
| 0.50 | 14.03 | 19.93 |
| 0.60 | 12.47 | 16.20 |
| 0.70 | 11.27 | 13.80 |
| 0.80 | 8.37 | 9.33 |
| 0.90 | 4.00 | 4.13 |

### Supplementary Figures

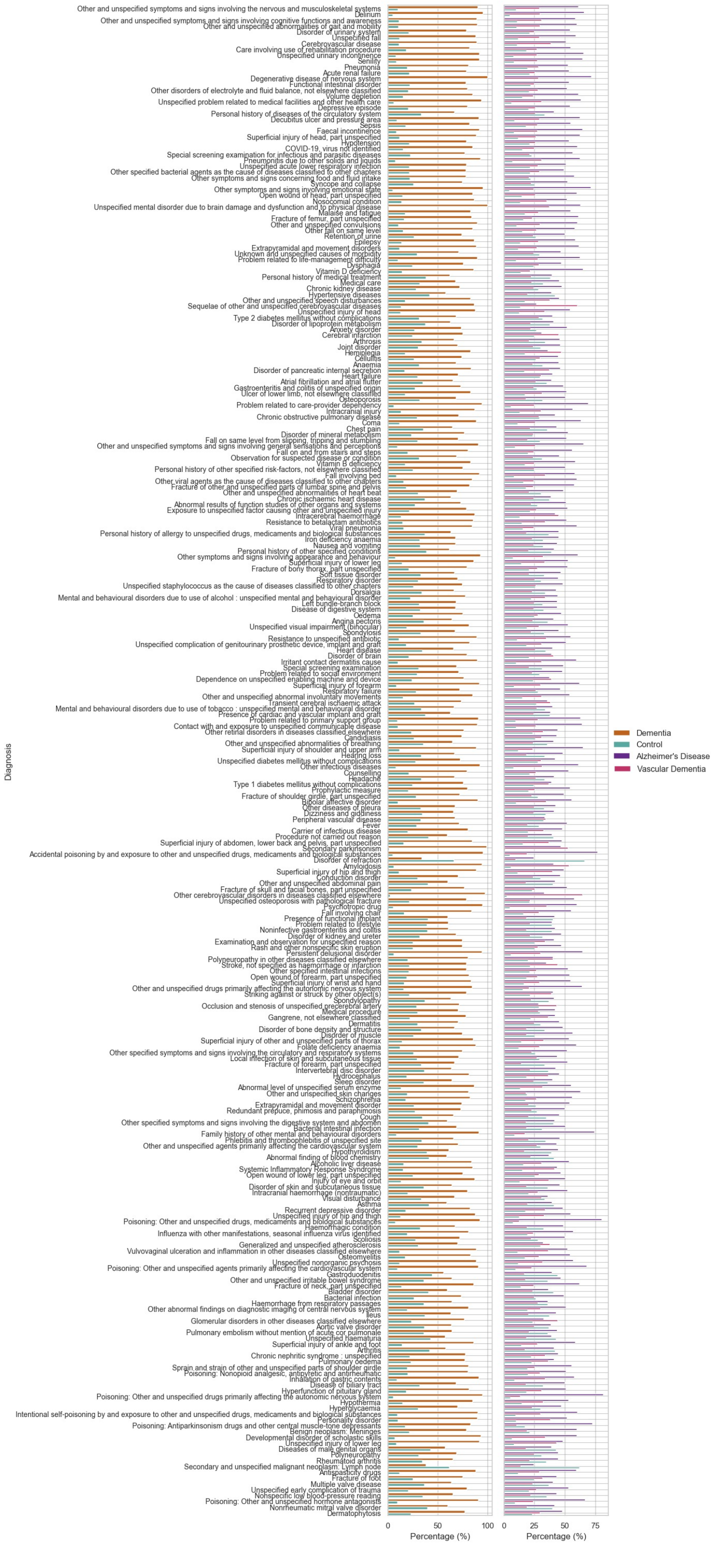


**Figure S1: Prevalence of key ICD-10 conditions (individual) that were significantly different between dementia and controls.** The first panel (left) is categorised by either controls or dementia cohort. The second panel (right) categorised by either controls or dementia subtypes Alzheimer's Disease and Vascular Dementia. Conditions are ordered from top to bottom, in order of most to least significant. Mann Whitney-U Test (all adjusted $p$-*value*s $<$ 0.05, Bonferroni correction) was applied.

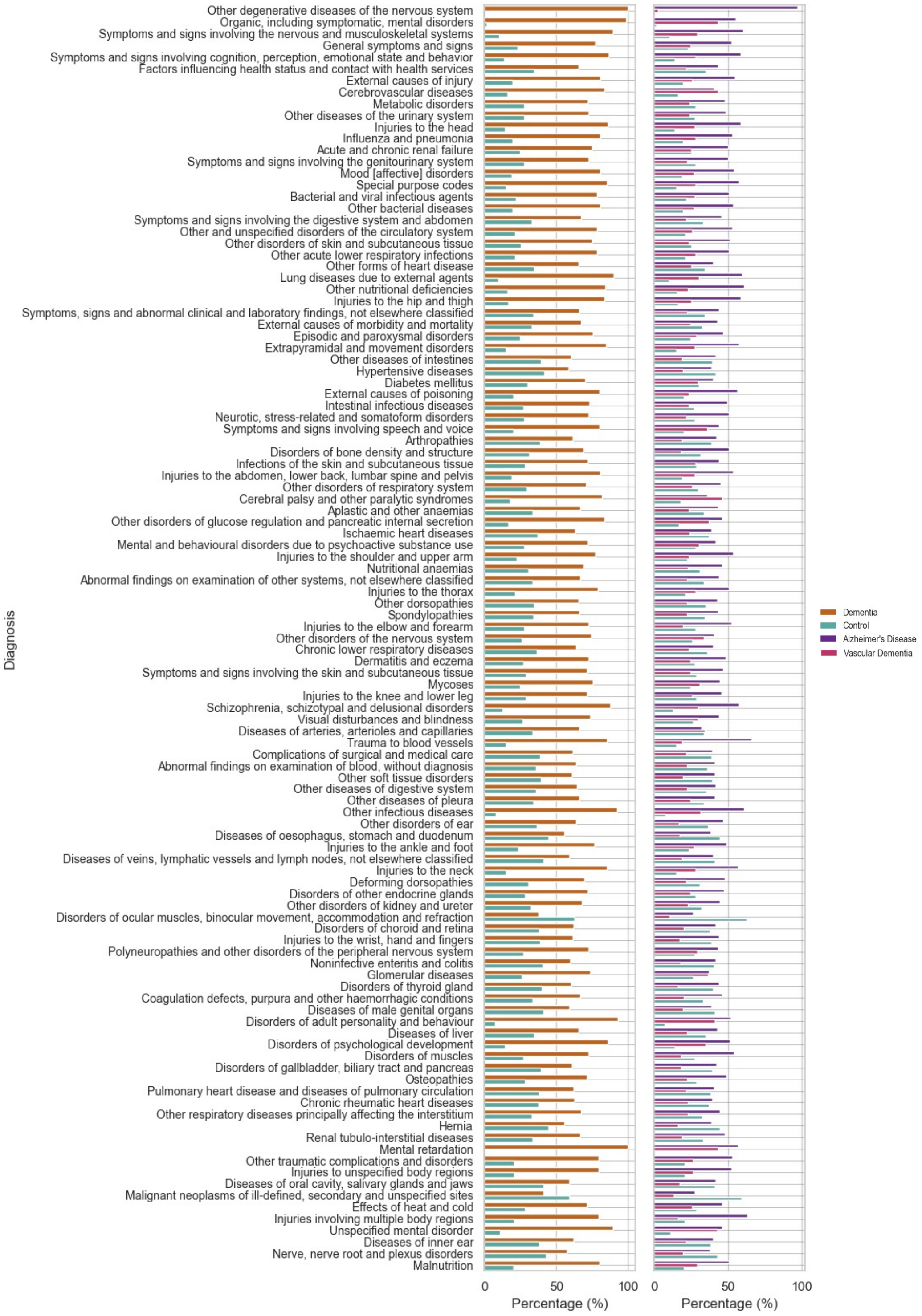


**Figure S2: Prevalence of key ICD-10 conditions (blocks) that were significantly different between dementia and controls.** The first panel (left) is categorised by either controls or dementia cohort. The second panel (right) categorised by either controls or dementia subtypes Alzheimer's Disease and Vascular Dementia. Conditions are ordered from top to bottom, in order of most to least significant. Circulatory, endocrine, bacterial/infectious, and intestinal diseases are all significantly different between the groups. Mann Whitney-U Test (all adjusted $p$-*value*s $< 0.05$, Bonferroni correction) was applied.

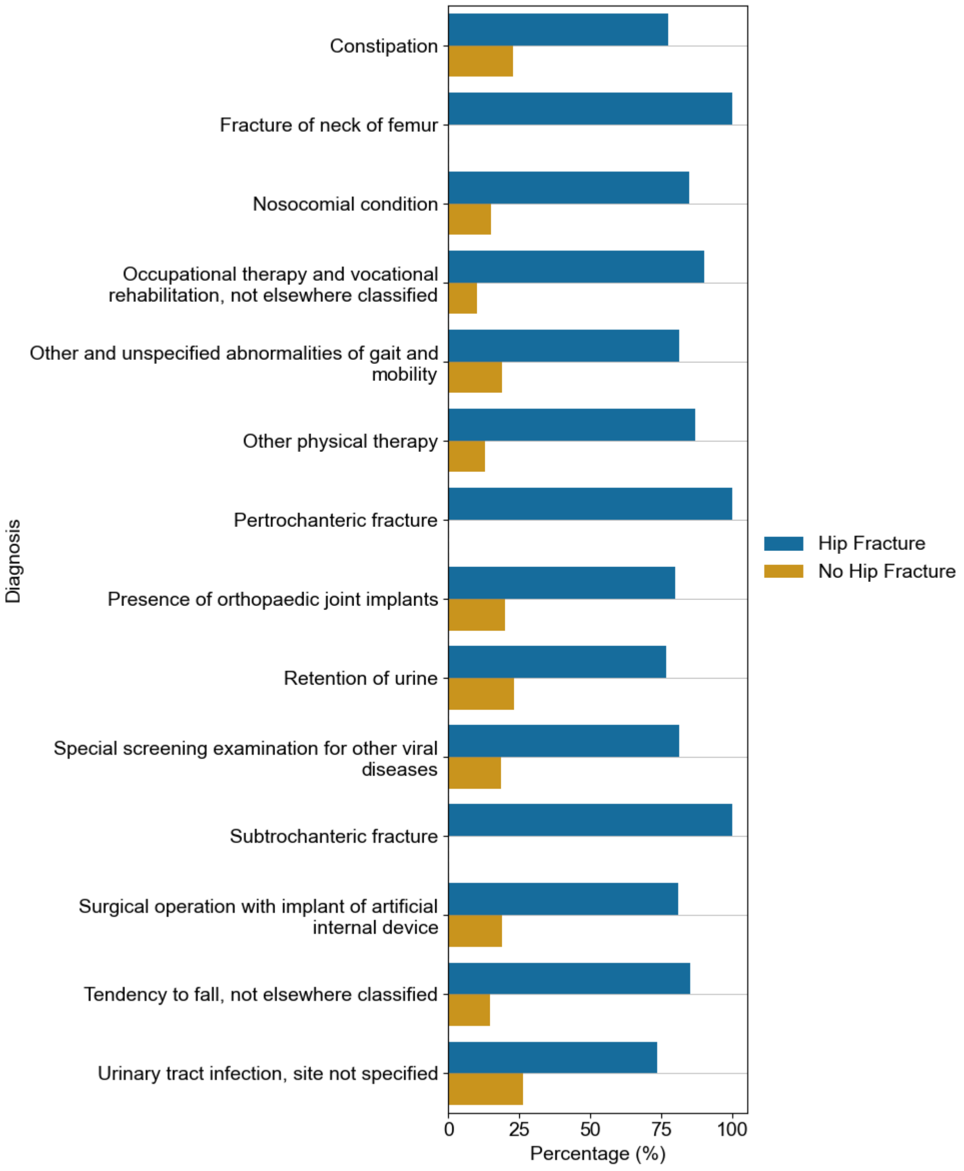


**Figure S3: Prevalence of key ICD-10 conditions (individual) that were significantly different between population of people with hip fractures and controls (population without hip fractures).** Conditions are ordered from top to bottom, in order of most to least significant. These conditions are not featured in the top 20 significant conditions, as with the dementia cohort and their matched controls. Mann Whitney-U Test (all adjusted $p$-*value*s $< 0.05$, Bonferroni correction) was applied.

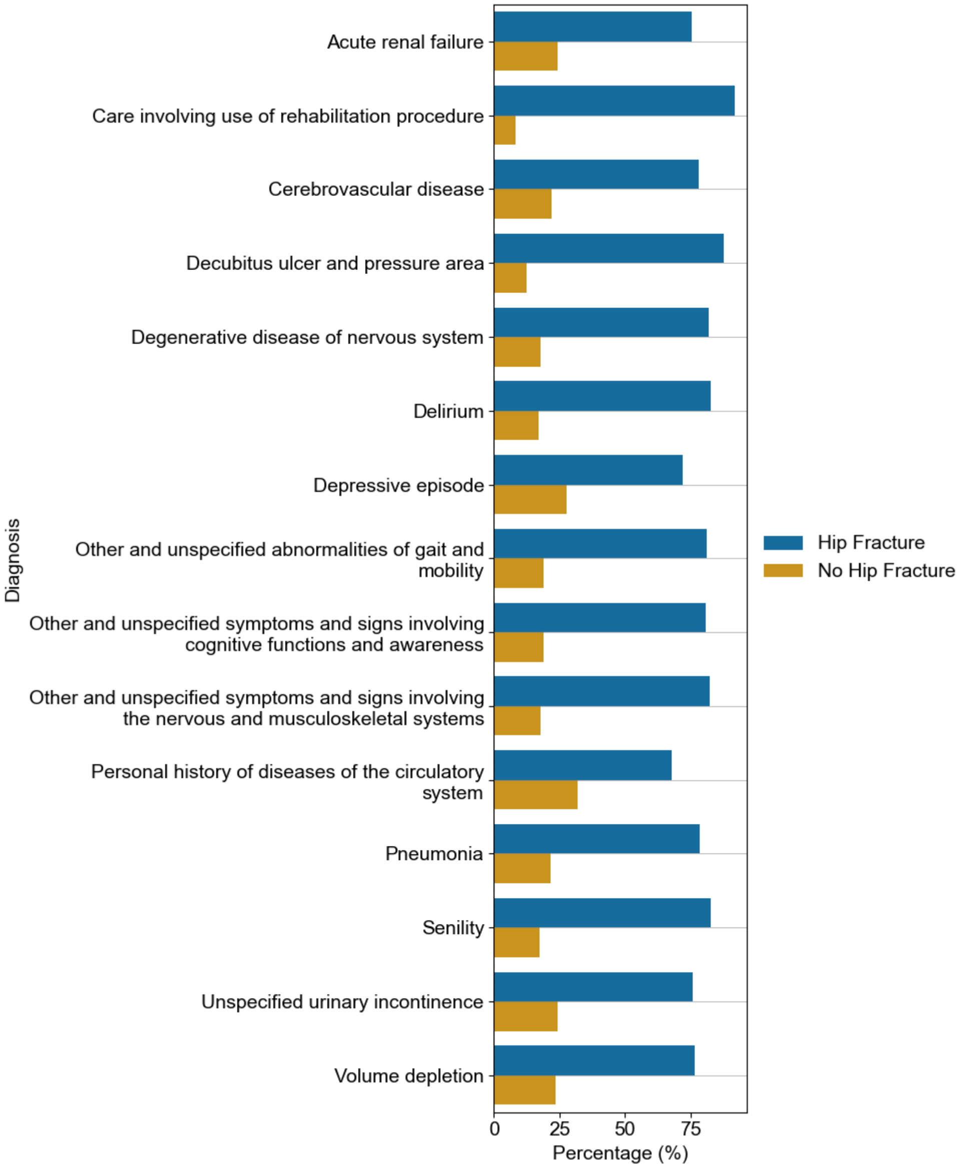


**Figure S4: Prevalence of key ICD-10 conditions (individual), in the hip fracture cohort, that were identified as conditions that were significantly different between dementia and controls.** Conditions are ordered from top to bottom, in order of most to least significant. The proportion of people with hip fractures are generally lower than what was seen in the dementia cohort. In addition, those without hip fractures have higher proportions than the matched controls in the dementia cohort. Mann Whitney-U Test (all adjusted $p$-*value*s $< 0.05$, Bonferroni correction) was applied.

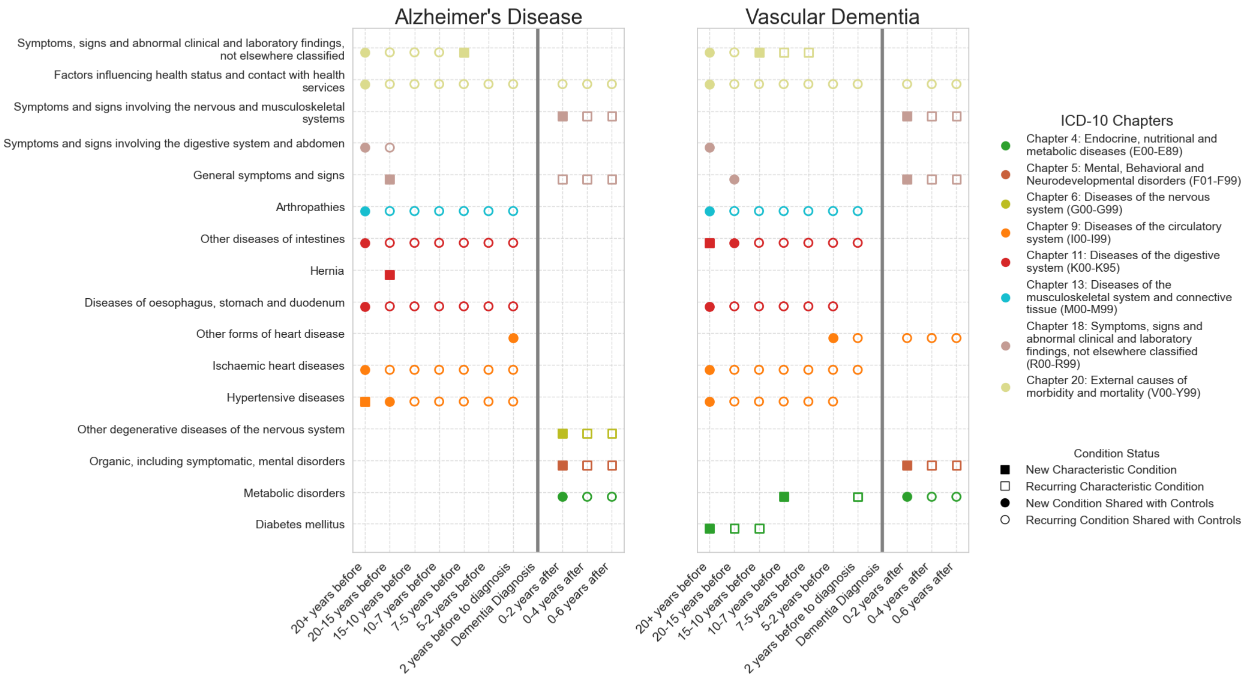


**Figure S5: Chronological mapping of ICD-10 blocks for comorbidities associated with Alzheimer's Disease (left panel) and Vascular Dementia (right panel), from 20 years before diagnosis to 6 years after.** Each time frame is shown, with the point of dementia diagnosis marked by a solid vertical grey line. The conditions displayed represent ICD-10 blocks or groups of comorbidities. As detailed in the first figure legend ('Condition Status'), squares represent ICD-10 blocks that were unique to the dementia sub-type cohort at the given time frame, while circles indicate blocks shared between the dementia and control cohorts. Filled shapes denote newly emerging blocks within the cohort, whereas unfilled shapes represent reoccurring blocks. Colours correspond to higher-level ICD-10 Chapters, coded in the 'ICD-10 Chapters' legend. (ICD: International Classification of Disease)

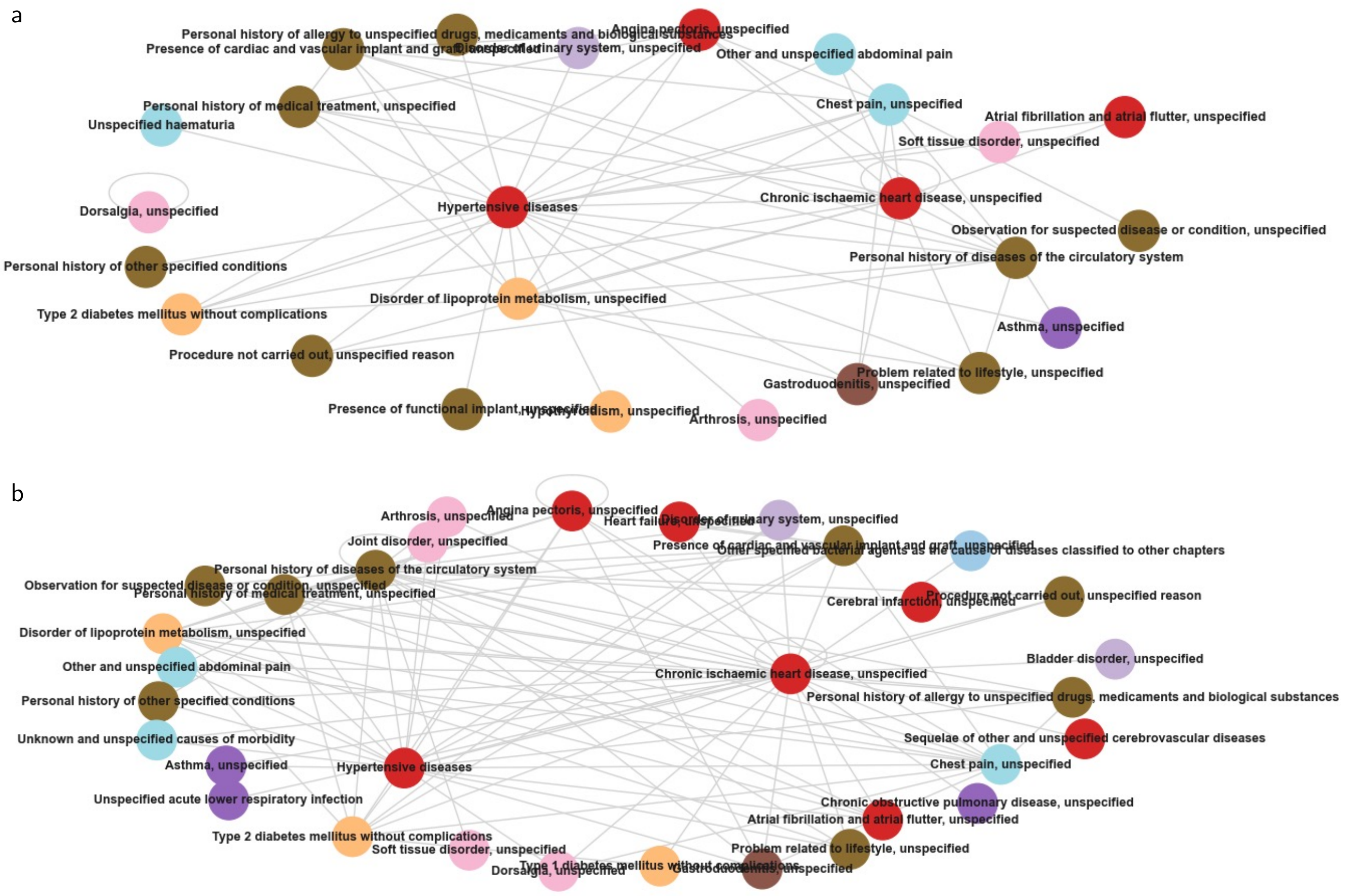


**Figure S6: Network analysis showing 15-10 years before diagnosis of Alzheimer's Disease and Vascular Dementia.** Panel (a) corresponds to network for Alzheimer's Disease cohort and panel (b) refers to the Vascular Dementia cohrot. Each node (ICD-10 condition) is coloured by the corresponding ICD-10 Chapter of that condition. The thickness of each edge (line between each condition) corresponds to how well-connected the condition is to other conditions in the time frame and sub-cohort. Conditions such as hypertension, circulatory disorders, diabetes and depression are common in both sub-types of dementia.

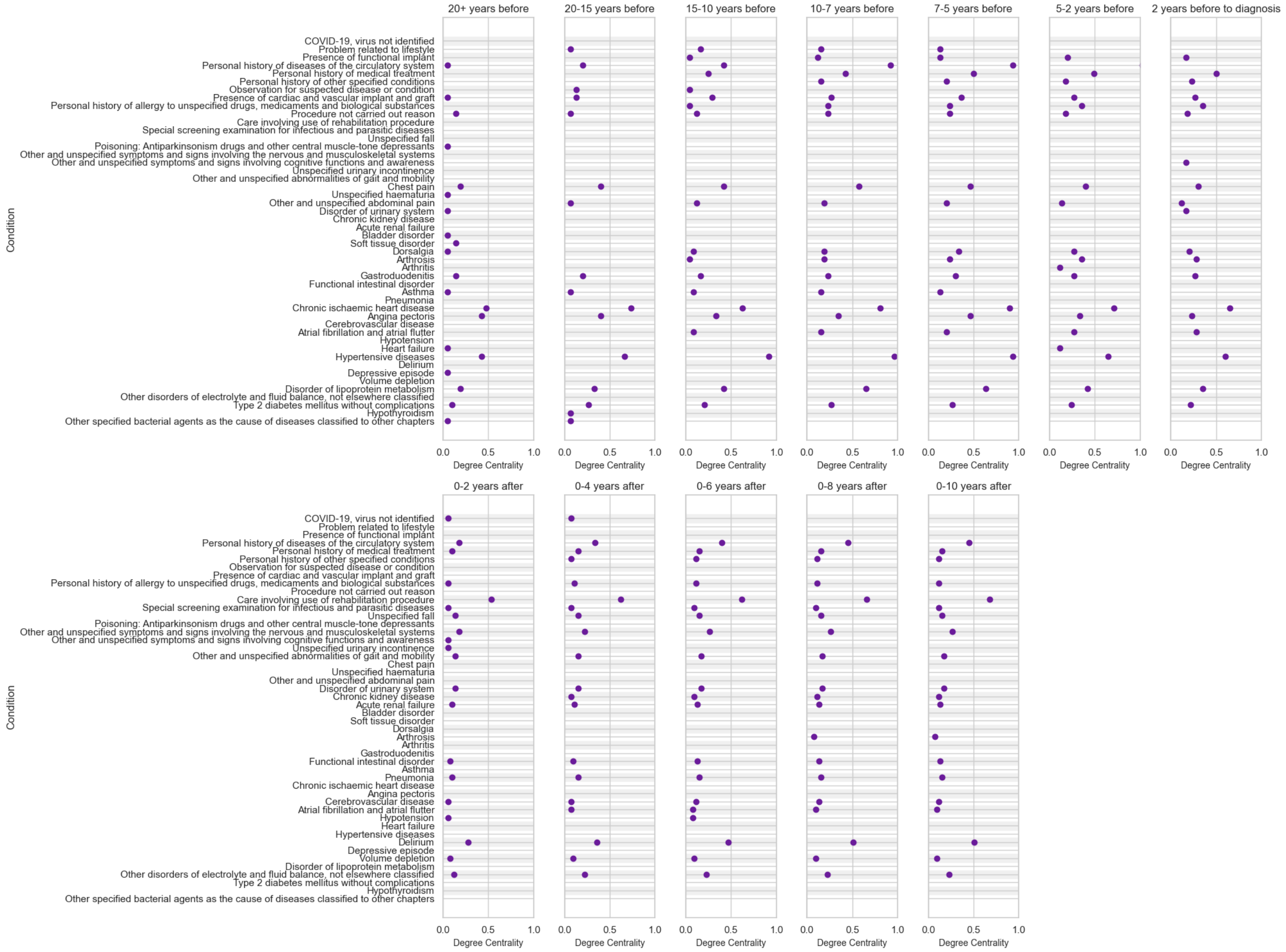


**Figure S7: ICD-10 conditions with the highest centrality measures, per time frame, as measured by undirected Bayesian Network Analysis, for Alzheimer's Disease cohort.** Centrality measures closer to 1.0 indicate the importance of that condition in relation to all other conditions. Hypertensive disease is seen to have high centrality at earlier time frames. Targeting conditions with higher centrality may have a more significant clinical impact.

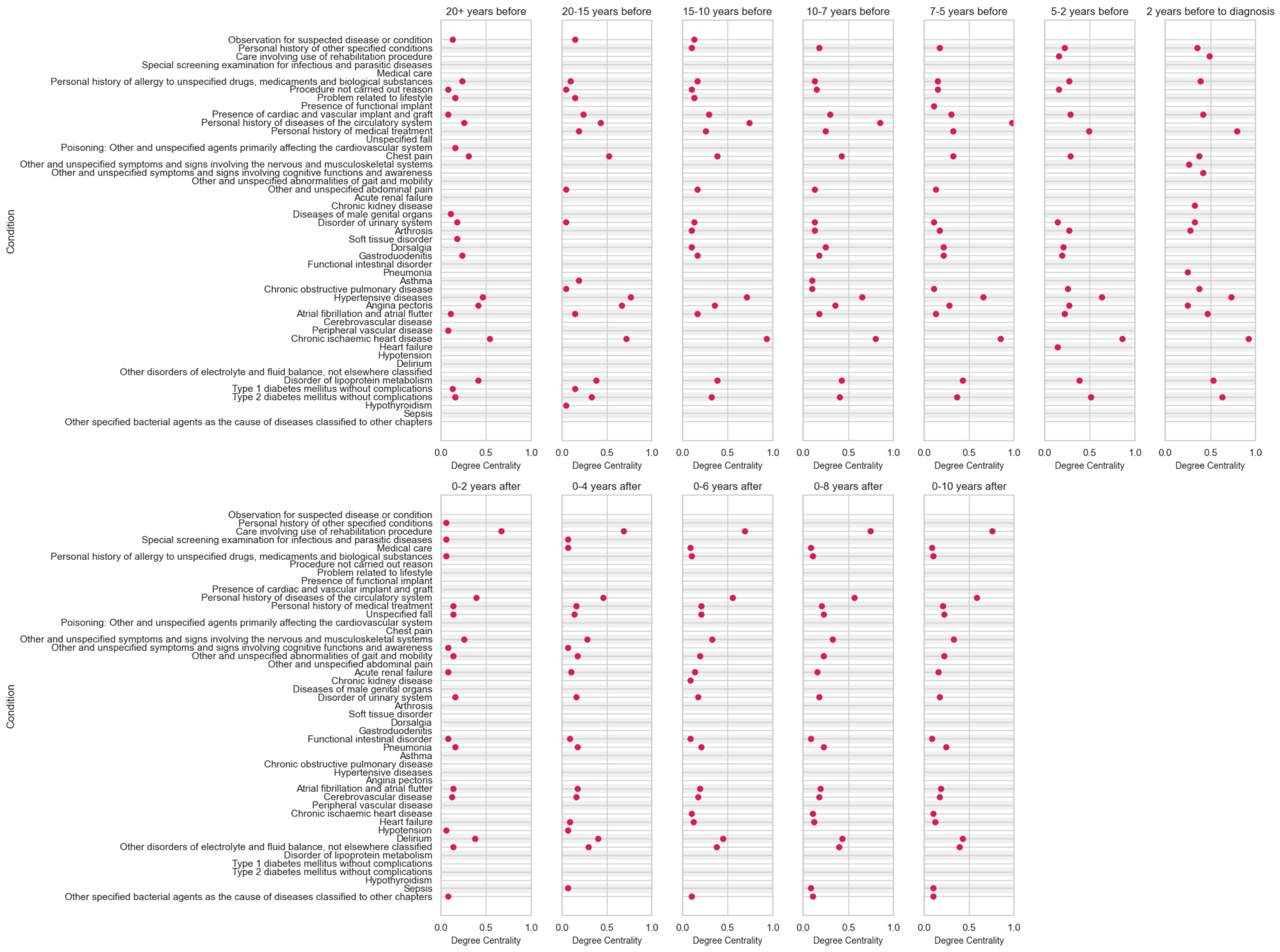

**Figure S8: ICD-10 conditions with the highest centrality measures, per time frame, as measured by undirected Bayesian Network Analysis, for Vascular Dementia cohort.** Centrality measures closer to 1.0 indicate the importance of that condition in relation to all other conditions. Chronic ischaemic heart disease and hypertensive disease are seen to have high centrality at earlier time frames. Targeting conditions with higher centrality may have a more significant clinical impact.
